# Epigenome-wide association studies identify novel DNA methylation sites associated with PTSD: A meta-analysis of 23 military and civilian cohorts

**DOI:** 10.1101/2024.07.15.24310422

**Authors:** Seyma Katrinli, Agaz H Wani, Adam X Maihofer, Andrew Ratanatharathorn, Nikolaos P Daskalakis, Janitza Montalvo-Ortiz, Diana L Núñez-Ríos, Anthony S Zannas, Xiang Zhao, Allison E Aiello, Allison E Ashley-Koch, Diana Avetyan, Dewleen G Baker, Jean C Beckham, Marco P Boks, Leslie A Brick, Evelyn Bromet, Frances A Champagne, Chia-Yen Chen, Shareefa Dalvie, Michelle F Dennis, Segun Fatumo, Catherine Fortier, Sandro Galea, Melanie E Garrett, Elbert Geuze, Gerald Grant, Michael A Hauser, Jasmeet P Hayes, Sian MJ Hemmings, Bertrand Russel Huber, Aarti Jajoo, Stefan Jansen, Ronald C Kessler, Nathan A Kimbrel, Anthony P King, Joel E Kleinman, Nastassja Koen, Karestan C Koenen, Pei-Fen Kuan, Israel Liberzon, Sarah D Linnstaedt, Adriana Lori, Benjamin J Luft, Jurjen J Luykx, Christine E Marx, Samuel A McLean, Divya Mehta, William Milberg, Mark W Miller, Mary S Mufford, Clarisse Musanabaganwa, Jean Mutabaruka, Leon Mutesa, Charles B Nemeroff, Nicole R Nugent, Holly K Orcutt, Xue-Jun Qin, Sheila A M Rauch, Kerry J Ressler, Victoria B Risbrough, Eugène Rutembesa, Bart P F Rutten, Soraya Seedat, Dan J Stein, Murray B Stein, Sylvanus Toikumo, Robert J Ursano, Annette Uwineza, Mieke H Verfaellie, Eric Vermetten, Christiaan H Vinkers, Erin B Ware, Derek E Wildman, Erika J Wolf, Ross McD Young, Ying Zhao, Leigh L van den Heuvel, PGC-PTSD Epigenetics Workgroup, PsychENCODE PTSD Brainomics Project, Traumatic Stress Brain Research Group, Monica Uddin, Caroline M Nievergelt, Alicia K Smith, Mark W Logue

## Abstract

**Background:** The occurrence of post-traumatic stress disorder (PTSD) following a traumatic event is associated with biological differences that can represent the susceptibility to PTSD, the impact of trauma, or the sequelae of PTSD itself. These effects include differences in DNA methylation (DNAm), an important form of epigenetic gene regulation, at multiple CpG loci across the genome. Moreover, these effects can be shared or specific to both central and peripheral tissues. Here, we aim to identify blood DNAm differences associated with PTSD and characterize the underlying biological mechanisms by examining the extent to which they mirror associations across multiple brain regions.

**Methods:** As the Psychiatric Genomics Consortium (PGC) PTSD Epigenetics Workgroup, we conducted the largest cross-sectional meta-analysis of epigenome-wide association studies (EWASs) of PTSD to date, involving 5077 participants (2156 PTSD cases and 2921 trauma-exposed controls) from 23 civilian and military studies. PTSD diagnosis assessments were harmonized following the standardized guidelines established by the PGC-PTSD Workgroup. DNAm was assayed from blood using either Illumina HumanMethylation450 or MethylationEPIC (850K) BeadChips. A common QC pipeline was applied. Within each cohort, DNA methylation was regressed on PTSD, sex (if applicable), age, blood cell proportions, and ancestry. An inverse variance-weighted meta-analysis was performed. We conducted replication analyses in tissue from multiple brain regions, neuronal nuclei, and a cellular model of prolonged stress.

**Results:** We identified 11 CpG sites associated with PTSD in the overall meta-analysis (1.44e-09 < *p* < 5.30e-08), as well as 14 associated in analyses of specific strata (military vs civilian cohort, sex, and ancestry), including CpGs in *AHRR* and *CDC42BPB*. Many of these loci exhibit blood-brain correlation in methylation levels and cross-tissue associations with PTSD in multiple brain regions. Methylation at most CpGs correlated with their annotated gene expression levels.

**Conclusions:** This study identifies 11 PTSD-associated CpGs, also leverages data from postmortem brain samples, GWAS, and genome-wide expression data to interpret the biology underlying these associations and prioritize genes whose regulation differs in those with PTSD.

## Introduction

Posttraumatic stress disorder (PTSD) is a serious psychiatric disorder characterized by intrusive memories of the traumatic event(s), avoidance of or numbing to situations that trigger those memories, and hyperarousal symptoms that can disturb mental and physical health (1).

These symptoms are associated with lower levels of self-care, lower compliance with medical treatment, and higher rates of substance use (2, 3). Thus, it is not surprising that PTSD increases the risk for chronic medical conditions, such as cardiovascular disorders, independent of lifestyle factors (e.g., substance use and sleep quality)(4, 5). Although most individuals experience at least one traumatic event, only a small fraction develop PTSD(6). Genetic and environmental factors contribute to this differential susceptibility in PTSD development upon trauma exposure (7, 8).

Genome-wide association studies (GWAS) of PTSD demonstrated remarkable success at identifying relevant genes, many of which are involved in the stress response or immune function (see reviews(9, 10)). The recent Psychiatric Genomics Consortium PTSD Workgroup (PGC-PTSD) Freeze 3 GWAS identified 95 genomic loci associated with PTSD, implicating genes involved in stress, immune, fear, and threat-related processes (11). Nonetheless, genetic differences do not fully account for an individual’s susceptibility to PTSD. Trauma exposure has been shown to alter epigenetic patterns in both animal and human studies, prompting the need to conduct epigenetic studies of PTSD in addition to genetic studies (12, 13). Epigenetic mechanisms are chemical modifications that can dictate the timing and magnitude of gene expression without altering the DNA sequence (14). The most widely studied epigenetic mechanism is DNA methylation (DNAm), which is defined as the addition of a methyl group to cytosine bases, particularly at cytosine-guanine dinucleotides (CpG sites). DNAm patterns respond to changes in the environment, are potentially reversible, and can be targeted for disease therapies (15, 16). Environmental influences on DNAm are apparent across the life span and may provide insight into the biological response to trauma(17).

Which specific DNAm sites differ across individuals and how they correlate with exposures and gene expression can vary across tissues (18). DNAm in human brain tissue, which is most relevant to the study of psychiatric disorders, is not easily accessible in living patients and hence is not a viable PTSD biomarker for clinical use. However, correlation has been observed between peripheral tissues (e.g., blood) and brain DNAm levels at specific genomic loci, and hence blood DNAm can potentially serve as a robust biomarker for implementing early intervention and developing improved preventative or therapeutic strategies for PTSD (19, 20).

Moreover, PTSD symptoms have been linked to the components of the peripheral immune system (21, 22) that can be readily assessed in blood DNAm. Multiple peripheral epigenome-wide association studies (EWASs) of PTSD identified CpGs in genes related to the immune system and neurotransmission(23–28). While prior EWASs of PTSD have reported promising results, the small sample sizes and variability of analysis methods across studies make it difficult to combine and interpret the findings effectively. Recent meta-analyses led by the PGC-PTSD Epigenetics Workgroup minimized these issues by increasing sample size, increasing sample diversity, and using a common quality control and analysis pipeline(29–33). These meta-analyses identified multiple new loci associated with PTSD, including *NRG1, AHRR, MAD1L1*, and *TBXAS1*, implicating immune dysregulation in those with PTSD (30–33).

Building on the prior work by Smith et al. (31), which conducted an EWAS meta-analysis in 1896 participants from 10 cohorts, this study includes 13 additional cohorts with a denser and more comprehensive DNAm array, bringing the sample up to 5077 participants from 23 civilian and military cohorts. Our current investigation replicated the findings of the initial PGC-PTSD epigenome-wide meta-analysis, reporting lower *AHRR* methylation in those with PTSD, and identified 8 new (11 total) PTSD-associated loci. We also leveraged data from postmortem brain samples, a cellular model of prolonged stress, GWAS, and genome-wide gene expression studies to interpret the biology underlying these associations and prioritize genes whose regulation differs in those with PTSD.

## Methods

### Cohorts and post-traumatic stress disorder assessments

The study includes 2156 current PTSD cases and 2921 trauma-exposed controls from 9 civilian cohorts: BEAR, DNHS, DCHS, GTP, NIU, Shared Roots, AURORA, H3A_Rwanda, WTC; and 9 military cohorts: GMRFQUT, MRS, PRISMO, Army STARRS, PROGrESS, NCPTSD/TRACTS, INTRuST, and VA cohorts (VA-M-AA & VA-M-EA). For DNHS, GTP, MRS, PRISMO, and Army STARRS, two different datasets were available based on the DNAm array. Two different datasets for these five cohorts did not have any overlapping samples and were treated as independent studies. Sample characteristics for the 23 studies that participated in the meta-analysis are summarized in Table 1.

**Table 1.**
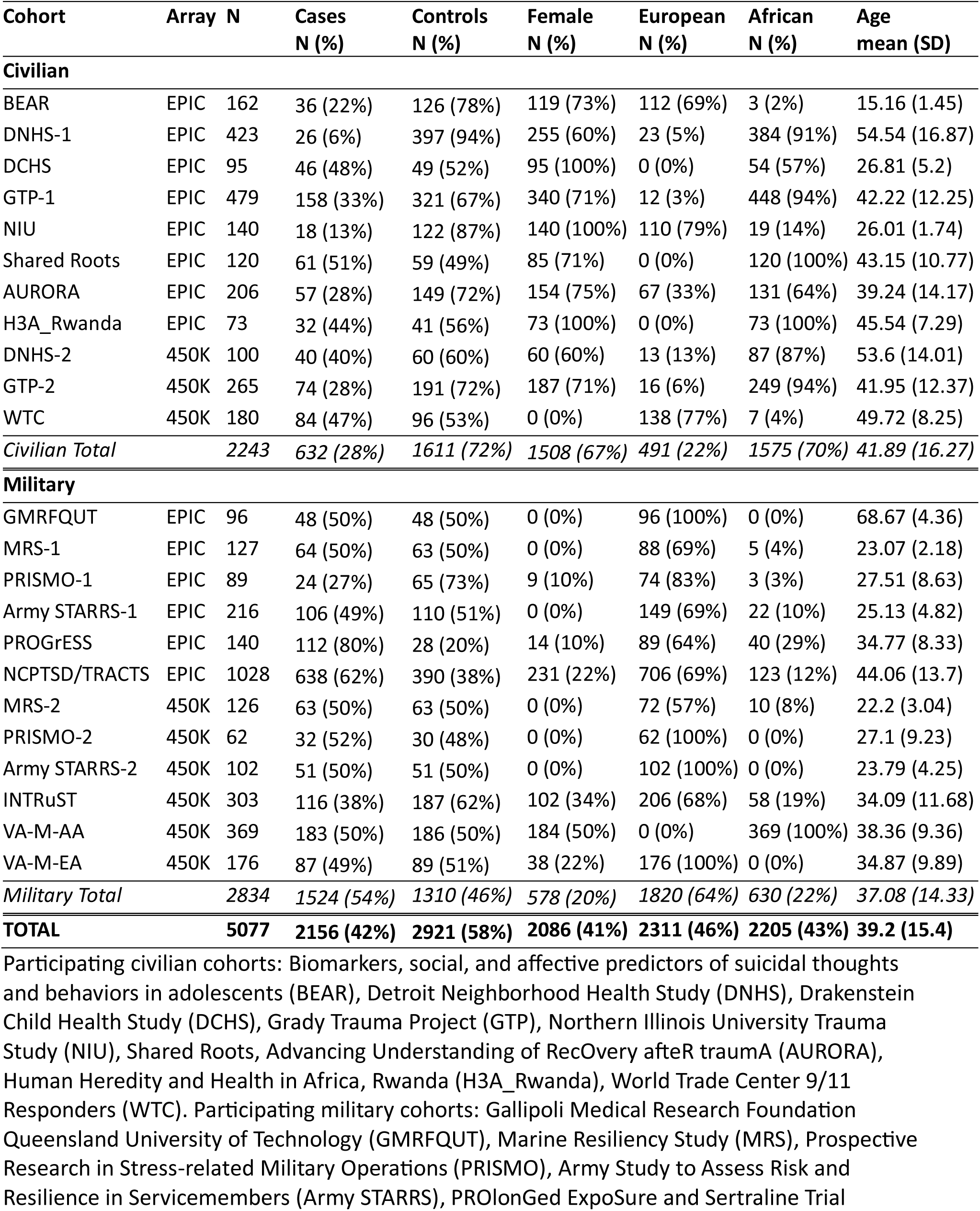
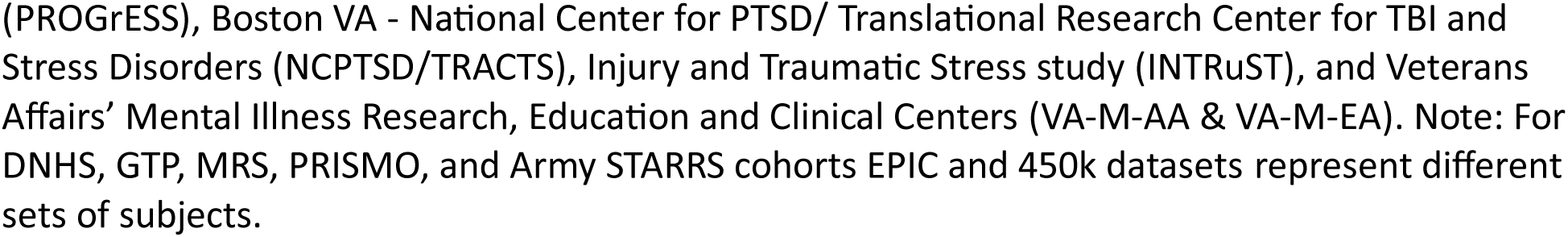
Overview of the studies.

| **Cohort** | **Array** | **N** | **Cases**<br>**N (%)** | **Controls**<br>**N (%)** | **Female**<br>**N (%)** | **European**<br>**N (%)** | **African**<br>**N (%)** | **Age**<br>**mean (SD)** |
| --- | --- | --- | --- | --- | --- | --- | --- | --- |
| **Civilian** |  |  |  |  |  |  |  |  |
| BEAR | EPIC | 162 | 36 (22%) | 126 (78%) | 119 (73%) | 112 (69%) | 3 (2%) | 15.16 (1.45) |
| DNHS-1 | EPIC | 423 | 26 (6%) | 397 (94%) | 255 (60%) | 23 (5%) | 384 (91%) | 54.54 (16.87) |
| DCHS | EPIC | 95 | 46 (48%) | 49 (52%) | 95 (100%) | 0 (0%) | 54 (57%) | 26.81 (5.2) |
| GTP-1 | EPIC | 479 | 158 (33%) | 321 (67%) | 340 (71%) | 12 (3%) | 448 (94%) | 42.22 (12.25) |
| NIU | EPIC | 140 | 18 (13%) | 122 (87%) | 140 (100%) | 110 (79%) | 19 (14%) | 26.01 (1.74) |
| Shared Roots | EPIC | 120 | 61 (51%) | 59 (49%) | 85 (71%) | 0 (0%) | 120 (100%) | 43.15 (10.77) |
| AURORA | EPIC | 206 | 57 (28%) | 149 (72%) | 154 (75%) | 67 (33%) | 131 (64%) | 39.24 (14.17) |
| H3A_Rwanda | EPIC | 73 | 32 (44%) | 41 (56%) | 73 (100%) | 0 (0%) | 73 (100%) | 45.54 (7.29) |
| DNHS-2 | 450K | 100 | 40 (40%) | 60 (60%) | 60 (60%) | 13 (13%) | 87 (87%) | 53.6 (14.01) |
| GTP-2 | 450K | 265 | 74 (28%) | 191 (72%) | 187 (71%) | 16 (6%) | 249 (94%) | 41.95 (12.37) |
| WTC | 450K | 180 | 84 (47%) | 96 (53%) | 0 (0%) | 138 (77%) | 7 (4%) | 49.72 (8.25) |
| *Civilian Total* |  | *2243* | *632 (28%)* | *1611 (72%)* | *1508 (67%)* | *491 (22%)* | *1575 (70%)* | *41.89 (16.27)* |
| **Military** |  |  |  |  |  |  |  |  |
| GMRFQUT | EPIC | 96 | 48 (50%) | 48 (50%) | 0 (0%) | 96 (100%) | 0 (0%) | 68.67 (4.36) |
| MRS-1 | EPIC | 127 | 64 (50%) | 63 (50%) | 0 (0%) | 88 (69%) | 5 (4%) | 23.07 (2.18) |
| PRISMO-1 | EPIC | 89 | 24 (27%) | 65 (73%) | 9 (10%) | 74 (83%) | 3 (3%) | 27.51 (8.63) |
| Army STARRS-1 | EPIC | 216 | 106 (49%) | 110 (51%) | 0 (0%) | 149 (69%) | 22 (10%) | 25.13 (4.82) |
| PROGrESS | EPIC | 140 | 112 (80%) | 28 (20%) | 14 (10%) | 89 (64%) | 40 (29%) | 34.77 (8.33) |
| NCPTSD/TRACTS | EPIC | 1028 | 638 (62%) | 390 (38%) | 231 (22%) | 706 (69%) | 123 (12%) | 44.06 (13.7) |
| MRS-2 | 450K | 126 | 63 (50%) | 63 (50%) | 0 (0%) | 72 (57%) | 10 (8%) | 22.2 (3.04) |
| PRISMO-2 | 450K | 62 | 32 (52%) | 30 (48%) | 0 (0%) | 62 (100%) | 0 (0%) | 27.1 (9.23) |
| Army STARRS-2 | 450K | 102 | 51 (50%) | 51 (50%) | 0 (0%) | 102 (100%) | 0 (0%) | 23.79 (4.25) |
| INTRuST | 450K | 303 | 116 (38%) | 187 (62%) | 102 (34%) | 206 (68%) | 58 (19%) | 34.09 (11.68) |
| VA-M-AA | 450K | 369 | 183 (50%) | 186 (50%) | 184 (50%) | 0 (0%) | 369 (100%) | 38.36 (9.36) |
| VA-M-EA | 450K | 176 | 87 (49%) | 89 (51%) | 38 (22%) | 176 (100%) | 0 (0%) | 34.87 (9.89) |
| *Military Total* |  | *2834* | *1524 (54%)* | *1310 (46%)* | *578 (20%)* | *1820 (64%)* | *630 (22%)* | *37.08 (14.33)* |
| **TOTAL** |  | **5077** | **2156 (42%)** | **2921 (58%)** | **2086 (41%)** | **2311 (46%)** | **2205 (43%)** | **39.2 (15.4)** |

Participating civilian cohorts: Biomarkers, social, and affective predictors of suicidal thoughts and behaviors in adolescents (BEAR), Detroit Neighborhood Health Study (DNHS), Drakenstein Child Health Study (DCHS), Grady Trauma Project (GTP), Northern Illinois University Trauma Study (NIU), Shared Roots, Advancing Understanding of RecOverY after trauma (AURORA), Human Heredity and Health in Africa, Rwanda (H3A\_Rwanda), World Trade Center 9/11 Responders (WTC). Participating military cohorts: Gallipoli Medical Research Foundation Queensland University of Technology (GMRFQUT), Marine Resiliency Study (MRS), Prospective Research in Stress-related Military Operations (PRISMO), Army Study to Assess Risk and Resilience in Servicemembers (Army STARRS), PROLonged ExpoSure and Sertraline Trial
(PROGrESS), Boston VA - National Center for PTSD/ Translational Research Center for TBI and Stress Disorders (NCPTSD/TRACTS), Injury and Traumatic Stress study (INTRuST), and Veterans Affairs' Mental Illness Research, Education and Clinical Centers (VA-M-AA & VA-M-EA). Note: For DNHS, GTP, MRS, PRISMO, and Army STARRS cohorts EPIC and 450k datasets represent different sets of subjects.

The sample is heterogeneous in terms of sex (41% female), ancestry (46% European, 43% African, and 11% of other ancestries), and cohort type (56% military cohort). Civilian cohorts skew towards being more female (67%) and African (70%), whereas military cohorts are predominantly male (80%) and European (64%).

All participants were exposed to a traumatic event and 42% met the criteria for current PTSD. The current PTSD diagnosis was assessed by each study following the standardized guidelines established by the PGC-PTSD Workgroup (7). Briefly, current PTSD diagnosis was determined based on the specific criteria set by the principal investigator of each study. All control subjects had experienced trauma. Subjects without a current PTSD but with a prior history of PTSD (i.e., remitted PTSD), were excluded. Detailed descriptions of cohorts and PTSD assessments are provided in the Supplement. All participants in these studies gave informed consent. The institutional review boards of each respective institution approved these studies.

### DNA methylation

Whole blood DNAm was measured using the Illumina MethylationEPIC BeadChip in 14 studies, and the HumanMethylation450 BeadChip in 9 studies (Table 1). All studies used a standardized consortium-developed quality control (QC) pipeline that differed somewhat depending on which chip was used. The 450K array pipeline (29, 31) is described in eMethods in the Supplement.

The EPIC pipeline (available at https://github.com/PGC-PTSD-EWAS/EPIC_QC) was similar to the 450K array pipeline. Samples with probe detection call rates lower than 90% and average intensity values that were either less than 50% of the overall sample mean or below 2000 arbitrary units (AU) were excluded. Probes with detection *p*-values > 0.01 were considered low quality and treated as missing. Probes that were missing in > 10% of the samples within the studies and were cross-hybridizing were removed (34). Data was normalized using single-sample Noob (ssNoob) implemented in R package *minfi* (*35*). *ComBat* was used to account for batch effects of chip and position while preserving PTSD, age, and sex effects (if applicable) (36). Blood-cell composition (i.e., the proportion of CD8+T, CD4+T, natural killer (NK), B cells, monocytes, and neutrophils) was estimated using the Robust Partial Correlation (RPC) method in *Epidish* (37) with a reference data specific to EPIC array (38). For studies without genome-wide genotype data (DNHS, NIU, Shared Roots, AURORA, H3A_Rwanda, GMRFQUT, PROGrESS), we estimated ancestry principal components (PCs) from DNAm data, using the method developed by Barfield et al. (39), as previously described (33). PCs 2 and 3, which were the components that correlate most with self-reported ancestry, were included as covariates (33, 39). In cohorts with available genome-wide genotype data, PCs 1-3 from GWAS were used to adjust for ancestry. We used R package *bacon* to control inflation, only if doing so results in the lambda being closer to 1 (40). To predict smoking status, a DNAm-based smoking score was calculated, as previously described (27) for cohorts with EPIC array data. A detailed description of DNAm-based ancestry PCs and smoking score calculation is provided in eMethods in the Supplement.

### Epigenome-wide association analysis

The association between PTSD and DNAm was tested using multivariable linear regression models within cohorts with balanced plate designs. For studies in which plate layouts were not balanced (Shared Roots, H3A_Rwanda, GMRFQUT), we conducted mixed-effect regression models, including chip as a random effect term. R package *CpGassoc* was used to fit the models (41). The models were adjusted for age, sex (if applicable), blood cell composition (i.e., CD8T, CD4T, NK, B cell, and monocyte cell proportions), and ancestry PCs. A post-hoc sensitivity analysis was performed by including a covariate for smoking: DNAm-based smoking score in studies with EPIC data and current smoking status for studies with 450K data. Furthermore, we conducted stratified analyses for both sexes, ancestry (European and African ancestry), and cohort type (civilian and military cohorts).

To combine results across studies, we performed inverse-variance weighting (IVW) meta-analysis in *meta* (42). Meta-analysis tested 411,786 CpGs common to 450K and EPIC arrays (23 studies), and 404,794 EPIC array specific CpGs (14 studies). Epigenome-wide significance threshold recommended for the MethylationEPIC BeadChip (*p* < 9.0E-08) was used to determine statistical significance (43). Gene Ontology (GO) enrichment analyses were conducted using the top 1000 CpGs in *missMethyl* (44). An FDR threshold of 5% was used to identify significant GO terms.

### Cross-tissue association analyses

#### Blood – Brain Correlations

The Blood Brain DNA Methylation Comparison Tool (19) was used to assess the correlations between methylation in blood and prefrontal cortex (PFC), entorhinal cortex (EC), superior temporal gyrus (STG), and cerebellum.

#### Postmortem brain DNAm

DNAm measured from post-mortem brains was obtained from two studies, each of which examined a unique but not necessarily distinct set of brain regions and cohorts: the National Center for PTSD Brain Bank cohort (NCPTSD-BB (45)) and the PsychENCODE Consortium for PTSD (PEC-PTSD) Brainomics cohort (46) (see eMethods in the Supplement for details), both of which were sourced from the Lieber Institute for Brain Development.

Methylation at PTSD-associated CpGs from the EWAS was tested for association with PTSD in DNA extracted from postmortem dorsolateral prefrontal cortex (dlPFC, BA9/46), ventromedial prefrontal cortex (vmPFC, BA12/32), amygdala, and dentate gyrus (DG). DNAm in the post-mortem tissue was measured using the EPIC array. We examined the associations with PTSD in dlPFC and vmPFC of 42 PTSD cases and 30 controls from the NCPTSD-BB. The associations between DNAm and PTSD in amygdala and DG were tested in 77 PTSD cases and 77 controls from the PEC-PTSD.

#### Neuronal nuclei

We examined cross-tissue association from neuronal nuclei isolation from the orbitofrontal cortex (OFC) of 25 PTSD cases and 13 healthy controls collected at the VA’s NCPTSD-BB (47). Fluorescence-Activated Nuclei Sorting (FANS) protocol was employed to isolate NeuN+ cells and the nuclei underwent reduced representation oxidative bisulfite-sequencing (RRoxBS), as previously described (48). We examined whether there was differential methylation and hydroxymethylation within 500 bp of CpGs from the epigenome-wide association analyses. Eight CpG sites match between the EPIC array and RRoxBS, two evaluating the same position (cg21566642 and cg25691167), and six within 500 bp (cg05575921, cg15148933, cg19558029, cg21161138, cg23576855, and cg26599989).

#### Cellular model of prolonged stress

We explored the associations between each of the PTSD-associated CpGs resulting from epigenome-wide association analyses and a cellular model of prolonged stress in which fibroblasts were subjected to physiological stress hormone (cortisol) levels for a prolonged period (51 days) as previously described (49, 50).

### Gene Regulation

Correlations between PTSD-associated CpG sites’ DNAm levels and expression levels of the corresponding gene (as determined by the EPIC array annotation) were tested in whole-blood RNA-sequencing (RNA-seq) data from participants in the BEAR (n=127), AURORA (n=173), NCPTSD Merit (n=204), and MRS cohorts (n=128 with multiple visits totaling 357 samples). The results were meta-analyzed using the IVW method using a Bonferroni correction for the number of CpGs examined. Detailed information about cohort-specific RNA-seq data generation is described in eMethods in the Supplement.

### Genetic effects

To evaluate the effect of nearby (<1 MB) polymorphisms on DNAm levels of CpGs associated with PTSD, we used cis-methylation quantitative trait locus (cis-meQTL) data from GoDMC(51) and meQTL EPIC(52) databases. For both databases, their default multiple testing adjustment was utilized: an FDR threshold of 5% in meQTL EPIC and *p* < 1e-08 in GoDMC. We checked the associations between the identified cis-SNPs and PTSD in the recent Freeze 3 GWAS from PGC-PTSD(11). Finally, we evaluated genetic associations between the genes with PTSD-associated DNAm changes and PTSD, using the gene-based test results from the recent PGC-PTSD Freeze 3 GWAS(11) using a Bonferroni correction for the 11 genes examined.

## Results

### Epigenome-wide association meta-analysis

We identified 11 PTSD-associated CpGs that passed the epigenome-wide significance threshold (*p* < 9e-08, Table 2, Figure 1, eFigure 1 in the Supplement). All CpG sites, except one site (cg21161138) near *AHRR*, remained nominally significant (1.52e-03 < *p* < 2.10e-07) with the same direction of association in the sensitivity analysis adjusted for smoking score (eTable 1 and eFigure 1 in the Supplement).

**Figure 1.**
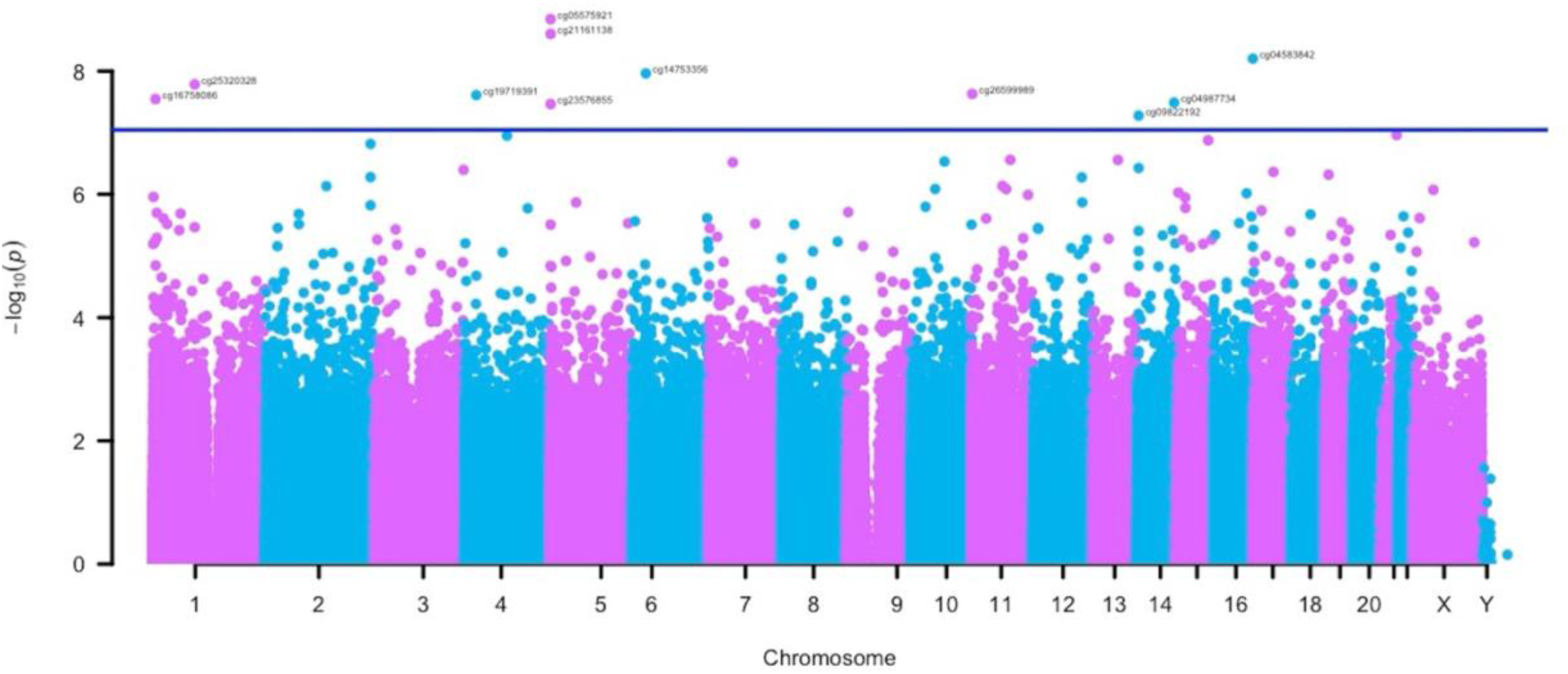
Manhattan Plot of the epigenome-wide association meta-analyses. The x-axis depicts chromosomes and the location of each CpG site across the genome. The y-axis depicts the -log10 of the unadjusted *p*-value for the association with current PTSD. Each dot represents a CpG site. The solid blue line indicates the epigenome-wide statistical significance at *p* < 9.0e-8.

**Table 2.**
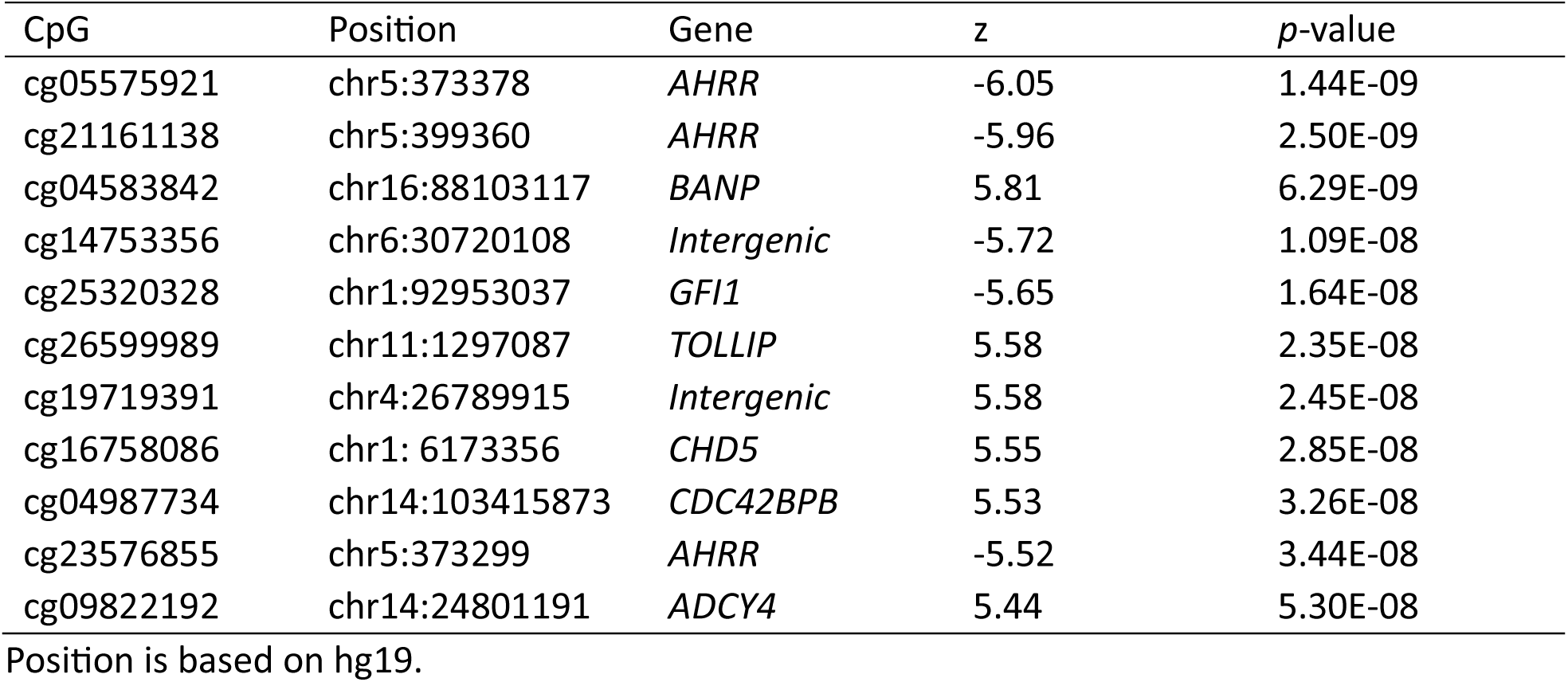
CpG sites associated with current PTSD in the primary meta-analysis.

### Stratified analyses for sex, ancestry, and cohort type

The directions of associations of the 11 significant CpGs were consistent at nominal significance (p < 0.05) when stratified by both sex, cohort type (civilian and military), and European ancestry. However, in the African ancestry stratified analysis, only 8 CpGs remained associated (p < 0.05) with PTSD (eTable 2 in the Supplement). We identified 1 epigenome-wide significant CpG site associated with PTSD in the female-stratified analysis, 1 in the male-stratified analysis, 5 in European ancestry-stratified analysis, 1 in the African ancestry-stratified analysis, 1 in the analysis for civilian-cohort analysis, and 5 in the military-cohort analysis (Table 3, eFigures 2-8 in the Supplement). Of note, some CpGs identified in the stratified analyses were specific to their stratum and were not associated with PTSD (*p* > 0.05) in the other stratum. For instance, cg25691167 in *FERD3L* was associated with PTSD in females (*p* = 4.24E-08), but not in males (*p* = 0.56) or the primarily male military cohorts (*p* = 0.13).

**Fig 2.**
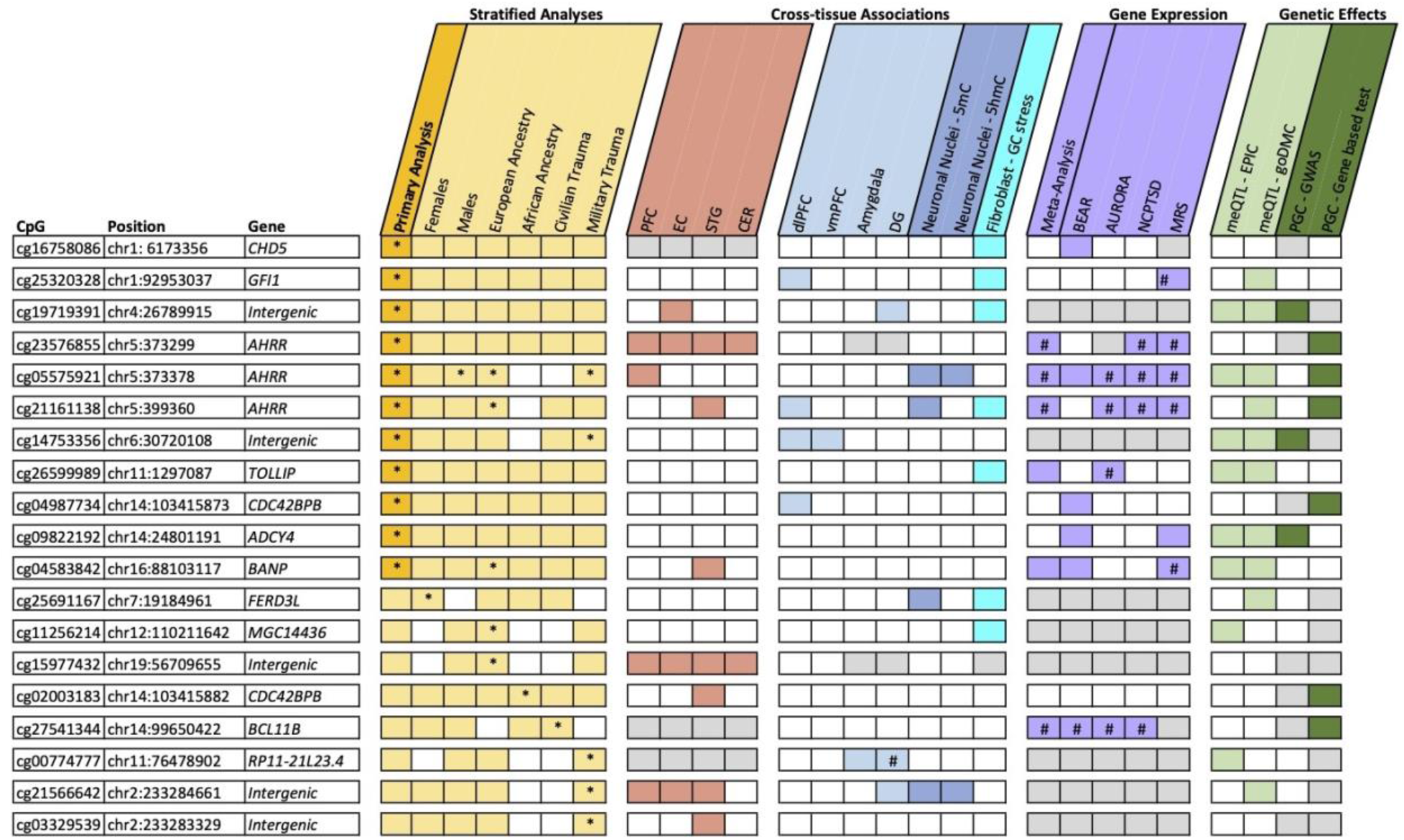
Summary of all analyses and findings. Figure combines CpGs from the main analysis (gold) and stratified analyses for sex, ancestry, and trauma type (light gold); and summarizes the results of blood and brain correlations (rose); cross-tissue associations for multiple brain regions (light blue), neuronal nuclei (blue), and a fibroblast model of prolonged stress (aqua), gene expression (purple); and genetic effects, including methylation quantitative trait loci (meQTL) analyses (light green) and genetic associations from the recent PGC-PTSD GWAS (dark green). Positive findings (*p* < 0.05) are indicated with the specific color of the respective category. Asterisk (*) indicates epigenome-wide significance (*p* < 9e-8). Gray represents the CpGs or genes were not present in the respective datasets. PFC: prefrontal cortex. EC: entorhinal cortex. STG: superior temporal gyrus. CER: cerebellum. dlPFC: dorsolateral prefrontal cortex. vmPFC: ventromedial prefrontal cortex. DG: dentate gyrus. 5mC: 5-Methylcytosine. 5hmC: 5-Hydroxymethylcytosine. GC: glucocorticoid.

**Table 3.** CpG sites associated with current PTSD in the stratified analyses.

| **CpG** | **Position** | **Gene** | **z** | **p-value** |
| --- | --- | --- | --- | --- |
| *Stratified analysis for females* |  |  |  |  |
| cg25691167 | chr7:19184961 | *FERD3L* | 5.48 | 4.24E-08 |
| *Stratified analysis for males* |  |  |  |  |
| cg05575921 | chr5:373378 | *AHRR* | -6.12 | 9.30E-10* |
| *Stratified analysis for European ancestry* |  |  |  |  |
| cg05575921 | chr5:373378 | *AHRR* | -6.85 | 7.24E-12* |
| cg11256214 | chr12:110211642 | *MGC14436* | 5.76 | 8.17E-09 |
| cg21161138 | chr5:399360 | *AHRR* | -5.62 | 1.95E-08* |
| cg15977432 | chr19:56709655 | *Intergenic* | -5.46 | 4.68E-08 |
| cg04583842 | chr16:88103117 | *BANP* | 5.43 | 5.66E-08* |
| *Stratified analysis for African ancestry* |  |  |  |  |
| cg02003183 | chr14:103415882 | *CDC42BPB* | 5.48 | 4.26E-08 |
| *Stratified analysis for civilian trauma* |  |  |  |  |
| cg27541344 | chr14:99650422 | *BCL11B* | 5.39 | 7.2073E-08 |
| *Stratified analysis for military trauma* |  |  |  |  |
| cg05575921 | chr5:373378 | *AHRR* | -6.22 | 4.92E-10* |
| cg00774777 | chr11:76478902 | *RP11-21L23.4* | 5.55 | 2.79E-08 |
| cg14753356 | chr6:30720108 | *Intergenic* | -5.47 | 4.53E-08* |
| cg21566642 | chr2:233284661 | *Intergenic* | -5.41 | 6.36E-08 |
| cg03329539 | chr2:233283329 | *Intergenic* | -5.36 | 8.53E-08 |

Position is based on hg19. The sites that were also epigenome-wide significant ( $p < 9\text{e-}08$ ) in the primary meta-analysis were indicated by an asterisk (\*).

### Gene enrichment analysis

We identified 3 GO term enrichments in the female-stratified analysis (FDR < 0.05), including nervous system development (eTable 3 in the Supplement). We did not identify any significant enrichments in the main analysis or other strata.

### Cross-tissue associations

Of the 19 significant CpGs from the primary and stratified analyses, 3 were not available in the Blood Brain DNA Methylation Comparison Tool database. Methylation at 9 of the remaining 16 CpGs was correlated between blood and at least one brain region (Figure 2, eTable4 in the Supplement). The strongest correlation was observed between the blood and PFC (r = 0.91) for cg23576855 (*AHRR*).

We next evaluated whether the PTSD-associated CpGs from blood were also associated with PTSD in the brain regions of dlPFC, vmPFC, amygdala, and DG, as well as neuronal nuclei. Out of 19 CpGs from the primary and stratified analyses in bulk tissue, 9 were nominally associated with PTSD in at least one brain region or the neuronal nuclei (*p* < 0.05, Figure 2, eTables 5 and 6 in the Supplement). For instance, PTSD cases had higher cg04987734 (*CDC42BPB*) methylation both in the blood (*p* = 3.26e-8) and the dlPFC (*p* = 3.9e-3). In addition, 7 CpGs exhibited nominally significant methylation changes (5 CpGs with the same direction of association) in fibroblasts when subjected to physiological stress hormone levels in a cellular model of prolonged stress (0.024 < ΔDNAm *<* 0.225*, p* < 0.05, Figure 2, eTable 7 in the Supplement)(49). Notably, the CpG sites (cg21161138 [*AHRR*], cg25320328 [*GFI*], cg19719391 [*intergenic*], cg25691167 [*FERD3L*]) that were consistently associated with PTSD in blood and postmortem brain also demonstrated a significant change in this cell model.

### Gene Regulation

Of the 11 PTSD-associated CpGs annotated to a gene, 10 correlated with their gene expression in at least one of the 4 RNA-seq cohorts (Figure 2, eTable 8 in the Supplement). Meta-analysis across cohorts identified 4 CpGs whose methylation levels were negatively correlated with their annotated gene expression after multiple test corrections: cg05575921, cg21161138, and cg23576855 with *AHRR*, and cg27541344 with *BCL11B* (p < 4.5e-03, -16.69 ≤ z ≤ -4.86 eTable 8 in the Supplement).

### Genetic regulation of PTSD-associated CpGs

Of the 19 CpGs from the primary and stratified analyses, 12 CpGs were associated with at least one nearby SNP within 1 MB according to GoDMC(51) and meQTL EPIC(53) databases (eTable 9 in the Supplement). In total, we identified 41 lead meQTLs, of which 8 were nominally associated with PTSD (*p* < 0.05, albeit in the opposite direction) in the PGC-PTSD Freeze 3 GWAS(11) (eTable 9 in the Supplement). For instance, lower methylation at the intergenic cg14753356 site is associated with PTSD (z = -5.72, *p* = 1.09E-08, Table 2) and higher methylation at cg14753356 is associated with rs28986310 T allele (beta = 0.31, *p* < 5e-324), which increases the risk of PTSD (z = 6.73, *p* = 1.68e-11, eTable 9 in the Supplement).

In addition, of the 11 genes harboring PTSD-associated CpGs, *AHRR*, *CDC42BPB*, and *BCL11B* were implicated in the recent PGC-PTSD gene-based analysis (3.48e-03 < *p* < 5.40e-05, eTable 10 in the Supplement). All genes are still significant after Bonferroni correction for 11 tests (p < 4.5e-03).

## Discussion

In the present epigenome-wide meta-analysis of blood DNAm levels, we identified 11 CpG sites associated with PTSD. Many of these are also associated with PTSD in multiple brain regions and a cellular model of prolonged stress exposure (49). Many of the PTSD-associated CpGs were associated with the expression of their respective genes. An examination of the most recent PGC-PTSD GWAS (11) indicated that many SNPs near significant CpGs were associated with both DNAm levels and PTSD diagnosis. Moreover, many of the EWAS-implicated genes were significant in gene-based tests from the GWAS. Thus, the GWAS results represent further evidence supporting the role of these genes in PTSD pathogenesis. However, the lower significance of the SNPs in association with PTSD relative to the corresponding CpGs (eTable 10 in the Supplement) indicates that the association between PTSD and the CpGs is likely not simply a byproduct of PTSD-associated SNPs.

Our *CDC42BPB* (CDC42 binding protein kinase beta) findings are of particular interest. *CDC42BPB* is involved in the regulation of cytoskeletal rearrangement, cell migration, and neurodevelopment(54). In the current study, increased *CDC42BPB* methylation at cg04987734 was associated with PTSD in blood and the dlPFC. Methylation at this site was also positively correlated with *CDC42BPB* expression in blood. Notably, higher methylation at cg04987734 has been associated with depressive symptoms (55) and increased C-reactive protein (CRP) levels (56, 57), which is perhaps not surprising given the bi-directional genetic association between PTSD and CRP levels (58). Multiple studies reported increased CRP levels and other inflammatory markers in those with PTSD, suggesting inflammation as an important component of PTSD (59–61). Future studies are warranted to investigate *CDC42BPB* methylation as a potential mediator of the relationship between PTSD and inflammation.

We observed 3 CpGs (cg05575921, cg21161138, and cg23576855) in *AHRR* (aryl-hydrocarbon receptor repressor) that were identified in the initial PGC-PTSD EWAS (31) and an independent study of US veterans (45). The aryl hydrocarbon receptor (AhR) plays a role in immunomodulation, including the regulation of T lymphocytes, B cell maturation, and the activity of macrophages, dendritic cells, and neutrophils (62), supporting the link between the immune system and PTSD. Although methylation at the *AHRR* CpGs is known to be influenced by smoking (63), cg05575921 and cg23576855 remained nominally significant (*p* = 1.52E-03 and p = 7.59E-04, respectively) in our sensitivity analysis adjusting for DNAm-based smoking scores. Notably, cg21161138 DNAm was also associated with PTSD in postmortem dlPFC and neuronal nuclei and demonstrated a significant change in the cellular model of prolonged stress (49), supporting the notion that there is an association between methylation at this locus and PTSD which is independent of the effects of smoking.

The stratified analyses identified DNAm-PTSD associations specific to sex, ancestry, and cohort type. The PTSD-associated site cg25691167 (*FERD3L*) in females (*p* = 4.24e-08) was not associated with PTSD in males (*p* = 0.57), suggesting that DNAm changes in cg25691167 might be sex-specific. Similarly, cg27541344 (*BCL11B*) was associated with PTSD in the civilian (*p* = 7.21e-08), but not the military cohorts (*p* = 0.38), whereas 3 out of 5 PTSD-associated CpGs in the military cohorts were not significant in the civilian cohorts (p > 0.05). We speculate that some of these DNAm changes specific to sex, ancestry, or cohort type might be due to unique characteristics of those strata, such as hormonal factors and environmental exposures.

## Strengths and Limitations

To our knowledge, this is the largest EWAS of PTSD to date. Our sample is diverse in terms of sex, ancestry, and cohort type. We leveraged data from postmortem brain samples, a cellular model of prolonged stress, GWAS, and genome-wide expression data to support our findings.

However, the study is not without limitations. First, methylation BeadChips only assess a subset of CpG sites in the genome; therefore, we may not capture all PTSD-associated CpG sites. Second, this is a cross-sectional study of participants with prior exposure to a traumatic event; thus, we were not able to assess whether the differences in DNAm between individuals with and without PTSD are a cause or consequence of PTSD or both. Third, our primary meta-analysis was performed using measures of blood DNAm. While this strategy provided valuable insights for future research on biomarkers of PTSD, it might not accurately represent the DNAm patterns within other tissues that are likely the most relevant to PTSD. However, the majority of the PTSD-associated CpG sites’ methylation levels were correlated between blood and at least one brain region. In addition, most PTSD-associated CpGs in blood were also associated with PTSD in one or more brain regions. Fourth, we used bulk tissue and adjusted for cellular heterogeneity, which might have obscured some signals, given the alterations in cell composition in those with PTSD(64, 65). Additionally, the brain regions examined varied between the online databases used to examine the blood-brain correlation of methylation values and the gene expression differences associated with PTSD, which can complicate interpretation. Finally, most cohorts that participated in the meta-analysis did not have detailed physical or psychiatric information on participants, including detailed information on trauma type and timing, PTSD symptom course, and treatment, making it challenging to evaluate and adjust for potential confounders, including substance use, comorbidities, or medication use.

## Conclusions

Taken together, this study replicates our previous findings and identifies novel PTSD-associated CpGs. Supporting data from multiple sources suggest that epigenetic mechanisms, particularly methylation in *AHRR* and *CDC42BPB*, may contribute to the complex relationship between the immune system and PTSD.

## Data Availability

The main summary statistics data that support the findings of this study will be available within Supplementary Data upon publication. Individual-level data from the cohorts or cohort-level summary statistics will be made available to researchers following an approved analysis proposal through the PGC-PTSD Epigenetics Workgroup with agreement of the cohort PIs. The raw data for the GTP cohort is available in the Gene Expression Omnibus database with the accession code GSE132203. Owing to military cohort data sharing restrictions, data from the VA, VA MIRECC, MRS, Army STARRS, PRISMO, PROGrESS, and NCPTSD/TRACTS cannot be publicly posted. However, such data can be provided in de-identified from a data repository through a data use agreement following applicable guidelines on data sharing and privacy protection. For additional information on access to these data, including PI contact information for the contributing cohorts, please contact the corresponding author.

## List of abbreviations

CpG: Cytosine-guanine dinucleotides
DG: Dentate gyrus
dlPFC: Dorsolateral prefrontal cortex
DNAm: DNA methylation
EWAS: Epigenome-wide association study
FANS: Fluorescence-Activated Nuclei Sorting
GWAS: Genome-wide association study
meQTL: Methylation quantitative trait locus
NK: Natural killer
OFC: Orbitofrontal cortex
PC: Principal component
PGC-PTSD: Psychiatric Genomics Consortium PTSD Workgroup
PTSD: Posttraumatic stress disorder
RPC: Robust Partial Correlation
RRoxBS: Reduced representation oxidative bisulfite-sequencing
vmPFC: Ventromedial prefrontal cortex

## Declarations

### Ethics approval and consent to participate

All participants in these studies gave informed consent. The institutional review boards of each respective institution approved these studies.

### Consent for publication

Not applicable.

### Competing interests

CYC is an employee of Biogen. NPD has served on scientific advisory boards for BioVie Pharma, Circular Genomics and Sentio Solutions for unrelated work. NRN serves as an unpaid member of the Ilumivu advisory board. SAMR receives support from the Wounded Warrior Project (WWP), Department of Veterans Affairs (VA), National Institute of Health (NIH), McCormick Foundation, Tonix Pharmaceuticals, Woodruff Foundation, and Department of Defense (DOD). Dr. Rauch receives royalties from Oxford University Press and American. KJR serves as a consultant for Acer, Bionomics, and Jazz Pharma; SABs for Sage, Boehringer Ingelheim, and Senseye. DJS has received consultancy honoraria from Discovery Vitality, Johnson & Johnson, Kanna, L’Oreal, Lundbeck, Orion, Sanofi, Servier, Takeda and Vistagen. MBS has in the past 3 years received consulting income from Acadia Pharmaceuticals, Aptinyx, atai Life Sciences, BigHealth, Biogen, Bionomics, BioXcel Therapeutics, Boehringer Ingelheim, Clexio, Delix Therapeutics, Eisai, EmpowerPharm, Engrail Therapeutics, Janssen, Jazz Pharmaceuticals, NeuroTrauma Sciences, PureTech Health, Sage Therapeutics, Sumitomo Pharma, and Roche/Genentech. MBS has stock options in Oxeia Biopharmaceuticals and EpiVario. MBS has been paid for his editorial work on Depression and Anxiety (Editor-in-Chief), Biological Psychiatry (Deputy Editor), and UpToDate (Co-Editor-in-Chief for Psychiatry). MBS has also received research support from NIH, Department of Veterans Affairs, and the Department of Defense. MBS is on the scientific advisory board for the Brain and Behavior Research Foundation and the Anxiety and Depression Association of America.

### Funding

This work was supported by the National Institute of Mental Health (NIMH; R01MH108826 and R01MH106595). This work was also supported by I01BX003477, a Department of Veterans Affairs BLR&D grant to MWL; 1R03AG051877, 1R21AG061367-01, RF1AG068121, and 1I01CX001276-01A2 to EJW; R21MH102834, 5I01CX000431, 5R01MH079806 to MWM; R01MD011728 to MU; R01MH105379 to NRN; R01MH117291, R01MH117292, R01MH117293 to FAC; R01MH108826 to NPD; U01MH115485 to LM, SF, SJ, JM, AU, and DEW; R01MH117291, R01MH117292, R01MH117293 to JEK; CDC/NIOSH U01OH012466 to PFK; R01MH117291, R01MH117292, R01MH117293 to CBN; R01MH093500 to CMN; 1R15MH099521-01, 5R21MH085436-02, 1R15HD049907-01A1 to HKO; R01MH108826 to AKS; I01BX002577, IK2CX000525, and lK6BX003777 to NAK; U01MH110925 to SAM; R21DA050160 and 1DP1DA058737 to JMO; Department of Veterans Affairs B9254-C to WM, B3001-C to CF; Department of Defense W81XWH-11-1-0073 and the National Center for Advancing Translational Sciences of the NIH UL1TR000433PEC-PTSD to SAMR; BX006186; BX005872 to VBR; VIDI award (91718336) from the Netherlands Scientific Organization to BPFR; Dutch Research Council (NWO) VIDI grant (09150171910042) to CHV. Brainomics work was supported by R01MH117291, R01MH117292, and R01MH117293. DJS and NK was supported by the South African Medical Research Council and the Bill and Melinda Gates Foundation (OPP 1017641). SS is supported by the South African Medical Research Council. SK is supported by the Building Interdisciplinary Research Careers in Women’s Health of the National Institutes of Health under Award Number K12HD085850.

### Author Contributions

PGC-PTSD writing group: S.K., M.W.L., C.M.N., A.K.S., and M.U.

Study PI or co-PI: A.E.A., A.E.A.-K., D.G.B., J.C.B., E.B., C.F., S.G., E.G., G.G., M.A.H., S.J., R.C.K., N.A.K., J.E.K., K.C.K., P.-F.K., M.W.L., B.L., C.E.M., S.A.M., W.M., M.W.M., L.M., C.B.N., C.M.N., N.R.N., H.K.O., S.A.M.R., K.J.R., V.R., S.S., A.K.S., D.J.S., M.B.S., M.U., R.J.U., E.V., D.E.W., E.J.W., and R.M.Y

Obtained funding for studies: J.C.B., M.P.B., S.F., C.F., E.G., M.A.H., R.C.K., K.C.K., M.W.L., J.J.L., S.A.M., M.W.M., C.M.N., N.R.N., H.K.O., S.A.M.R., K.J.R., B.P.F.R., A.K.S., M.U., R.J.U., and E.V.

Clinical: D.G.B., J.C.B., M.F.D., S.F., C.F., E.G., J.P.H., N.A.K., N.K., C.M., J.M., S.A.M.R., K.J.R., E.R., A.U., M.H.V., E.V., E.J.W., and L.L.V.D.H.

Contributed data: A.E.A.-K., D.G.B., J.C.B., F.A.C., N.P.D., M.F.D., C.F., M.A.H., J.P.H., S.M.J.H., B.R.H., S.J., S.K., N.A.K., A.P.K., K.C.K., P.-F.K., I.L., A.L, B.L., J.J.L., D.M., W.M., M.W.M., J.M.-O., C.M., L.M., C.M.N., N.R.N., S.A.M.R., V.R., S.S., A.K.S., M.B.S., S.T., M.U., A.U., M.H.V., D.E.W., E.J.W., A.S.Z., and L.L.V.D.H.

Statistical analysis: A.E.A.-K., D.A., M.P.B., L.B., C.-Y.C., S.D., N.P.D., S.F., M.E.G., A.J., S.K., A.P.K., I.L., M.W.L., A.X.M., D.M., M.S.M., C.M.N., D.L.N.-R., X-J.Q., A.R., B.P.F.R., A.K.S., C.H.V., A.H.W., E.B.W., A.S.Z., X.Z., and Y.Z.

Bioinformatics: M.P.B., L.B., C.-Y.C., N.P.D., M.E.G., M.A.H., A.J., S.K., S.D.L., M.W.L., A.X.M., A.R., B.P.F.R., A.H.W., X.Z., and Y.Z.

Genomics: M.P.B., M.A.H., S.D.L., A.X.M., B.P.F.R., C.H.V., A.S.Z., and X.Z. PI of the EWAS group: M.W.L., C.M.N., A.K.S., and M.U.

## Acknowledgments

PGC-PTSD Epigenetics Workgroup: Reid S Alisch, Ananda B Amstadter, Don Armstrong, Archana Basu, Jean C Beckham, Nicole L Bjorklund, Barbara H Chaiyachati, Judith B M Ensink, Segun Fatumo, Leland L Fleming, Sandro Galea, Joel Gelernter, Ryan J Herringa, Sonia Jain, Diana L Juvinao-Quintero, Seyma Katrinli, Elizabeth Ketema, José J Martínez-Magaña, Burook Misganaw, Shiela Tiemi Nagamatsu, Danny M Nispeling, John Pfeiffer, Christian Schmahl, Gen Shinozaki, Clara Snijders, Jennifer A Sumner, Patricia C Swart, Audrey Tyrka, Robert J Ursano, Mirjam van Zuiden, Eric Vermetten, Jaqueline S Womersley, Nagy A Youssef, Yuanchao Zheng, Yiwen Zhu, Lea Zillich PsychENCODE PTSD Brainomics Project: Dhivya Arasappan, Sabina Berretta, Rahul A. Bharadwaj, Frances A. Champagne, Leonardo Collado-Torres, Christos Chatzinakos, Nikolaos P. Daskalakis, Chris P. DiPietro, Duc M. Duong, Amy Deep-Soboslay, Nick Eagles, Louise Huuki, Thomas Hyde, Artemis Iatrou, Aarti Jajoo, Joel E. Kleinman, Charles B. Nemeroff, Geo Pertea, Deanna Ross, Nicholas T. Seyfried, Joo Heon Shin, Kerry J.Ressler, Clara Snijders, Ran Tao, Daniel R. Weinberger, Stefan Wuchty, Dennis Wylie Traumatic Stress Brain Research Group: Victor E. Alvarez, David Benedek, Alicia Che, Dianne A. Cruz, David A. Davis, Matthew J. Girgenti, Ellen Hoffman, Paul E. Holtzheimer, Bertrand R. Huber, Alfred Kaye, John H. Krystal, Adam T. Labadorf, Terence M. Keane, Ann McKee, Brian Marx, Crystal Noller, Meghan Pierce, William K. Scott, Paula Schnurr, Krista DiSano, Thor Stein, Douglas E. Williamson, Keith A. Young

## Notes

### Author Declarations

IRBs of each respective institution (Lifespan Hospitals, University of Michigan, University of North Carolina at Chapel Hill, University of Cape Town,Stellenbosch University,the Western Cape Provincial Health Research, Emory University, Northern Illinois University, University Teaching Hospital of Kigali, College of Medicine and Health Sciences at the University of Rwanda, Stony Brook University, the Queensland University of Technology, Greenslopes Private Hospital, University of California San Diego, VA San Diego Research Service, Naval Health Research Center, University Medical Center Utrecht, Uniformed Services University of the Health Sciences for the Henry M. Jackson Foundation, the Institute for Social Research at the University of Michigan, VHAAAHS, VASDHCS, CHSVAMC, MGH, the Department of Defense Human Research Protection Office, Boston VA Healthcare System, Salisbury VA, Hampton VA, Durham VA and Duke University Medical Centers) gave ethical approval for this work.

