## Supplemental Files for "Epigenome-wide association studies identify novel DNA methylation sites associated with PTSD: A meta-analysis of 23 military and civilian cohorts"

### Supplementary Information

**eMethods.** Supplementary Methods
**eFigure 1.** Forest Plots of PTSD-associated CpGs in each cohort.
**eFigure 2.** Manhattan plots of the stratified analyses.
**eFigure 3.** Forest Plots of CpGs associated with PTSD in the stratified analysis for females.
**eFigure 4.** Forest Plots of CpGs associated with PTSD in the stratified analysis for males.
**eFigure 5.** Forest Plots of CpGs associated with PTSD in the stratified analysis for European ancestry.
**eFigure 6.** Forest Plots of CpGs associated with PTSD in the stratified analysis for African ancestry.
**eFigure 7.** Forest Plots of CpGs associated with PTSD in the stratified analysis for civilian trauma.
**eFigure 8.** Forest Plots of CpGs associated with PTSD in the stratified analysis for military trauma.
**eTable 1.** PTSD-associated CpGs from the primary analysis in the sensitivity analysis adjusted for smoking score.
**eTable 2.** PTSD-associated CpGs from the primary analysis in the stratified analysis for sex, ancestry, and trauma type
**eTable 3.** Gene Ontology Enrichment results in females.
**eTable 4.** Correlation values of methylation between blood and brain regions from Blood Brain DNA Methylation Comparison Tool.
**eTable 5.** Results of PTSD-associated CpGs from blood in the brain.
**eTable 6.** Results of PTSD-associated CpGs from blood in neuronal nuclei.
**eTable 7.** Results of PTSD-associated CpGs from blood in fibroblasts.
**eTable 8.** DNA methylation and cis-gene expression (RNAseq) correlations.
**eTable 9.** Significant SNPs from cis-meQTL analysis and their association with PTSD.
**eTable 10.** Genetic associations of the identified PTSD-associated genes from gene-based tests.
**References.**

**eMethods**

**Cohort Information**

*Biomarkers, social, and affective predictors of suicidal thoughts and behaviors in adolescents (BEAR).* The BEAR study, described in detail elsewhere[1], involved a sample of 194 adolescents (ages 13-18) who had been hospitalized for suicidal thoughts/behaviors. Blood collection and clinical interview procedures were completed during hospitalization. Trauma exposure and symptoms of posttraumatic stress disorder (PTSD) were assessed via clinical interview with gold standard measures, specifically the Life Events Checklist for DSM-5[2] and the Clinician Administered PTSD Scale for Children and Adolescents for DSM-5[3]. If participants endorsed multiple traumatic experiences, participants were asked to focus on current symptoms related to the most upsetting event. All participants were consented to the present research and procedures were approved by the Lifespan Hospitals IRB.

*Detroit Neighborhood Health Study (DNHS).* As detailed in a previous study[4], participants (n=1,547 at the baseline wave) were assessed for PTSD symptoms using the PTSD checklist (PCL-C), a 17-item self-report measure of Diagnostic and Statistical Manual of Mental Disorders (DSM-IV) symptoms, and additional questions about duration, timing, and impairment or disability due to the symptoms[5]. Participants were initially asked to identify Potentially Traumatic Events that they experienced in the past from a list of 19 events. PTSD symptoms were then assessed by referencing two traumatic events that the respondent may have experienced: one that the participant regarded as the worst and one randomly selected event from the remaining PTEs a respondent may have experienced. Respondents were considered affected by lifetime PTSD if all six DSM-IV criteria were met in reference to either the worst or the random event. Current PTSD was then identified based on whether participants endorsed experiencing symptoms within the last month. The DNHS was approved by the institutional review board at the University of Michigan and University of North Carolina at Chapel Hill.

*Drakenstein Child Health Study (DCHS).* The Drakenstein Child Health Study (DCHS)[6], is a population based birth cohort, with participants initially recruited in a peri-urban area in South Africa. Pregnant women were enrolled from March 2012 to March 2015. Pregnant women who were 18 years or older, between 20-28 weeks gestation, and residing in the area were enrolled from 2 public health clinics. Study visits were synchronized with health care and immunization visits. During the first two years of the study, additional study visits occurred at 6, 12 and 24 months, and mother-infant pairs could also participate in intensive biweekly follow-up during the first year. Mother-child pairs were followed from birth through childhood, collecting longitudinal measurements of risk factors encompassing environmental, nutritional, immunological, genetic, infectious, and maternal factors. Maternal smoking or passive smoke exposure was self-reported. A composite measure of socioeconomic status (SES), encompassing current employment, education, household income and an asset index, was based on previous work in the South African Stress and Health Study. Maternal mental health measures included assessments of PTSD, depression, psychological distress, and intimate partner violence (IPV) both antenatally and postnatally. For the purpose of this study, PTSD was assessed using The Mini International Neuropsychiatric Interview (MINI)[7,8].

*Grady Trauma Project (GTP).* GTP research participants were approached in the waiting rooms of the primary care clinic or obstetrical-gynecological clinic of a large, urban, public hospital in Atlanta, GA while either waiting for their medical appointments or while waiting with others who were scheduled for medical appointments[9]. Subjects willing to participate provided written informed consent and participated in a verbal interview and blood draw. Current and lifetime PTSD diagnosis was assessed by clinical psychologists using the Clinician-Administered PTSD Scale for DSM IV (CAPS-4)[10,11] or the modified PTSD Symptomatic Scale (PSS)[12]. For this study, cases were specified as having current PTSD, and controls had no current or lifetime history of the disorder. Demographic variables including age, sex and race were assessed through self-report. The Institutional Review Boards of Emory University School of Medicine and the Research Oversight Committee of Grady Memorial Hospital approved this study.

*Northern Illinois University Trauma Study (NIU).* Data were collected from women who completed multiple waves of a federally-funded longitudinal study (NIH 5R21MH085436-02) approved by the Northern Illinois University (NIU) Institutional Review Board. Between August 2006 and February 14, 2008, female students enrolled in an Introductory Psychology course received partial course credit for completing an initial assessment for a longitudinal study. Participants provided written informed consent and completed computerized surveys in individual face-to-face sessions. Following the NIU campus shooting on February 14$^{th}$, 2008 (the fourth deadliest university shooting in U.S, history at that time), the NIU Trauma Study was launched to examine risk and protective factors following the event. Women previously enrolled in the ongoing longitudinal investigation were recruited to participate in a multi-wave longitudinal study examining post-shooting adjustment. Seventeen days following the shooting, eligible participants (*n* = 812 of the 885 participants who consented to follow-up contact and were current NIU students) were invited to participate in the post-shooting adjustment study via e-mail. Of these participants, 691 (85.10%) completed the initial post-shooting assessment and were recruited for subsequent post-shooting assessments over the next 30 months. Between October 2012 and February 2015, these 691participants were recruited via e-mail, phone call, and mail contact to participate in a federally-funded study (NIH 1R15MH099521-01) including a fear-potentiated startle experimental session. Of those eligible, 131 (18.96%) completed the experimental session and provided blood and saliva samples. Written informed consent was obtained for the experimental session and samples. The Institutional Review Board of Northern Illinois University approved this study. Current PTSD diagnosis at the first post-shooting assessment was assessed by self-report on the Distressing Events Questionnaire (DEQ)[13] and cases were specified as having a score greater or equal to 18 on the DEQ and controls had a score of 17 or lower. Demographic variables including age, race, and ethnicity were assessed through self-report.

*Shared Roots.* SHRS or "Understanding the SHARED ROOTS of Neuropsychiatric Disorders and Modifiable Risk Factors for Cardiovascular Disease" was a matched case-control study examining the factors that contribute to the comorbidity of metabolic syndrome and neuropsychiatric disorders[14]. Controls were trauma-exposed. Potentially traumatic events were identified using the Life Events Checklist for DSM-5 (LEC-5)[2]. The CAPS-5[15] was administered by clinicians to assess PTSD over the prior month. The CAPS-5 and the PTSD Checklist for DSM-5 (PCL-5)[16] were both administered to assess PTSD symptom severity. Lifetime diagnosis of PTSD was not assessed for in this study. Childhood trauma was assessed using the Childhood Trauma Questionnaire (CTQ)[17]. Demographic variables including age, sex and race were assessed through self-report. The Institutional Review Board of Stellenbosch University approved this study (HREC: N13/08/115).

*Advancing Understanding of RecOvery afteR traumA (AURORA).* The AURORA study is a large multi-ancestry cohort study of women and men presenting to the ED within 72 hours after exposure to psychological trauma[18]. Inclusion and exclusion criteria for AURORA participants were as follows. Patients aged 18–75 years who presented to the ED within 72 hours of trauma exposure at participating emergency department (ED) sites were screened for study eligibility. Trauma exposures automatically qualifying for study enrollment were motor vehicle collision, physical assault, sexual assault, fall greater than 10 feet, or mass casualty incidents. Other trauma exposures also qualified if: (1) the individual responded to a screener question that they experienced the exposure as involving actual or threatened serious injury, sexual violence, or death, either by direct exposure, witnessing, or learning about it; and (2) the research assistant agreed that the exposure was a plausible qualifying event. Exclusion criteria included administration of general anesthesia, long bone fractures, laceration with significant hemorrhage, solid organ injury > American Association for the Surgery of Trauma Grade 1, not alert and oriented at the time of enrollment, not fluent in written or spoken English, visual or auditory impairment precluding completion of web-based neurocognitive evaluations and/or telephone follow-ups, self-inflicted or occupational injury, prisoners, individuals pregnant or breastfeeding, individuals reporting ongoing domestic violence, and individuals taking > 20 mg morphine or equivalent per day. Eligible patients also needed to have an iOS or Android-compatible smartphone with internet access and an email address that they check regularly. Probable PTSD diagnosis at 6 months was defined using the PTSD Checklist for DSM-5 (PCL-5) – a 20-item self-report scale that uses a 0-4 response format asking how much the participant was “bothered by” each PTSD symptom (0-4 scale) in the past 30 days – and a previously established PCL-5 score threshold of ≥31[19-21]. The present study is focused on a subset of the AURORA cohort with available phenotypic, DNA methylation, and RNA sequencing data at the 6-month follow-up after trauma exposure (n=206, 57 (27.675%) current probable PTSD, 154 (74.76%) women, 67 (32.52%) White, 131 (63.59%) Black. With a mean age of 39.55 (SD=14.34)).

*Human Heredity and Health in Africa, Rwanda (H3A_Rwanda).* Data from the H3A_Rwanda cohort were obtained from a subset of participants in a previously published pilot study (n=30)[22,23] and from newly recruited participants via support from the H3Africa Consortium (n=40)[24]. All study participants were women of Tutsi ethnicity who were pregnant during the genocide. The pilot study participants were recruited via genocide survivor’s association known as Association of Genocide Widows Agahozo (AVEGA) and PTSD was assessed by a trained psychologist according to DSM-IV criteria using the PCL-17[25], translated into Kinyarwanda. The

H3Africa-affiliated participants were recruited in collaboration with local NGOs who work closely with the target population, mainly the AVEGA and Compassion International Rwanda, a religiously inspired organization that helps children in the community. The data collection was conducted via 2019-2023 and PTSD was assessed via a self-administered questionnaire using the PCL-5 including criterion A[26], translated into Kinyarwanda, according to DSM 5 criteria. This tool was validated by Harvard Trauma Questionnaires (HTQ). All subjects provided written informed consent and participated in a blood draw. Participants in the pilot study were recruited prior to those in the H3Africa-related study. Two participants overlapped between studies, such that among the 73 samples included in the current meta-analysis there were 71 unique participants. One of the two repeat participants was a control at both time points, whereas the other participant was a PTSD case at one time point and a control at another. The two studies were approved by the Institutional Review Board of the University Teaching Hospital of Kigali (pilot study- Ref EC/CHUK/025/11) and The Institutional Review Board of the College of Medicine and Health Sciences at the University of Rwanda (No.370/CMHS/IRB/2020).

*World Trade Center Responders (WTC)*. As described by Bromet and colleagues[27,28], responders enrolled in the Stony Brook University/Long Island WTC Health Program were administered the SCID PTSD module modified to assess PTSD in relation to WTC exposures. The sample providing blood samples for the epigenetics assays was restricted to men (the vast majority of the responders) and oversampled for posttraumatic stress disorder (PTSD)[29]. The Committees on Research Involving Human Subjects at Stony Brook University approved the study.

*Gallipoli Medical Research Foundation Queensland University of Technology (GMRFQUT).* The GMRF-QUT cohort comprised of a large cohort of veterans who have been or were currently being treated for PTSD at the Keith Payne Unit within the Greenslopes Private Hospital in Queensland, Australia[30]. Subjects willing to participate provided written informed consent and participated in a verbal interview with a clinician and blood draw. The Clinician Administered PTSD Scale for DSM 5 (CAPS-5)[10,11] was used to assess PTSD over the prior 2 weeks and lifetime by clinical psychologists. Respondents were considered to have a current diagnosis if CAPS-5 criteria were met. For this study, cases were specified as having current PTSD, and controls had no current or lifetime history of the disorder. Demographic variables including age, sex and race were assessed through self-report questionnaires. Ethics approval for the project was obtained from the Human Research Ethics Committee of the Queensland University of Technology and Greenslopes Private Hospital. This study was carried out in accordance with The Code of Ethics of the World Medical Association (Declaration of Helsinki).

*Marine Resiliency Study (MRS).* In the MRS[31,32], PTSD was diagnosed up to 3 times, once before deployment and 3 and/or 6 month post deployment. Post-traumatic stress (PTS) symptoms were assessed using a structured diagnostic interview, the Clinician Administered PTSD Scale (CAPS), and PTSD diagnosis followed the DSM-IV criteria for partial and full PTSD and a minimum CAPS score of 40. None of the participants in the EWAS had PTSD at pre-deployment (CAPS scores $\leq 25$). Samples of PTSD cases were selected from the 3- or 6-months post-deployment visits depending on which visit had the highest CAPS score. Combat-exposed controls with low to no PTSD-symptoms were selected from post-deployment visits, matching for age, ancestry, and time of post-deployment visit. The study was approved by the institutional review boards of the University of California San Diego, VA San Diego Research Service, and Naval Health Research Center. All participants provided informed consent.

*Prospective Research in Stress-related Military Operations (PRISMO)*. PRISMO is a large prospective study of well-characterized Dutch military soldiers scheduled for a deployment of at least four months to Afghanistan with longitudinal follow-up[33,34]. Current PTSD symptoms were assessed using the Self-Report Inventory for PTSD (SRIP)[35], and blood samples were collected approximately 1 month before and at both 1 and 6 months after deployment. Traumatic stress exposure during deployment to Afghanistan was assessed with a 19-item deployment experiences checklist, as previously reported[36]. PTSD cases were selected for this DNA methylation study from the 1- or 6-months post-deployment visits, depending on which visit had the highest SRIP score. All controls were combat-exposed and had low PTSD symptoms[37]. The study was approved by the Institutional Review Board of the University Medical Center Utrecht (Utrecht, the Netherlands), and was conducted in accordance with the Declaration of Helsinki. Written consent was obtained from all participants.

*The Army Study to Assess Risk and Resilience in Servicemembers (Army STARRS)*. Army STARRS is a multicomponent prospective study of risk-resilience factors for suicidality and its psychopathological correlates among US Army personnel[38]. Participants completed a computerized version of the Composite International Diagnostic Interview screening scales (CIDI-SC) and a 6-item screening version of the PTSD Checklist (PCL-6) for DSM-IV ~6 weeks before deployment to Afghanistan, and the PCL for DSM-IV at 1-, 2-, and 6-months post-deployment. Trauma exposure was assessed from answers pertaining to childhood, adulthood civilian, and military traumatic events. PTSD diagnosis was assigned using multiple imputation methods that relied on PCL and CIDI-SC data[39]. Whole blood for methylation assays was collected approximately 6 weeks before deployment and 1-month post-deployment. None of the subjects were diagnosed with PTSD prior to deployment. PTSD cases were selected based on their PTSD diagnosis at 6 months post-deployment. Controls were subjects without PTSD, matched on age, deployment stress, and childhood adversity. The recruitment, consent, human subject, and data protection procedures were approved by the Human Subjects Committees of the Uniformed Services University of the Health Sciences for the Henry M. Jackson Foundation (the primary grantee), the Institute for Social Research at the University of Michigan (the organization collecting the data), and all other collaborating organizations. All subjects provided informed consent.

*PROlonGed ExpoSure and Sertraline Trial (PROGrESS)*. The participants included in the study were part of a larger study of the PROlonGed ExpoSure and Sertraline Trial (PROGrESS), a four-site [VA Ann Arbor Healthcare System (VAAAHS), Ralph H. Johnson VA Medical Center (CHSVAMC), Massachusetts General Hospital (MGH), and VA San Diego Healthcare System (VASDHCS) randomized-controlled trial (RCT; N = 223), designed to examine: 1) the comparative effectiveness of Prolonged Exposure plus placebo (PE/PLB), Sertraline plus Enhanced Medication Management (SERT/EMM), or combined treatment (PE/SERT) on PTSD, and 2) neurobiological predictors and potential biomarkers of treatment response including hypothalamic-pituitary-adrenal axis (HPA), brain, and genetic/genomic biomarkers[40,41]. PROGrESS was approved by the institutional review boards at VHAAAHS, the University of Michigan, VASDHCS, CHSVAMC, MGH and the Department of Defense Human Research Protection Office (HRPO). Participants provided written informed consent before enrollment. Participants and clinicians were blinded to pill condition through week 24, and independent evaluators were blinded to treatment assignments for the duration of the study. The PROGrESS study methods were published in detail[40,41]. Briefly, inclusion criteria were service members or veterans of the Iraq or Afghanistan wars with combat-related PTSD and significant impairment (Clinicians-Administered PTSD Scale for DSM-IV [CAPS-IV] [26] score ≥ 50) of at least 3 months' duration. Exclusion criteria included factors related to safety and appropriateness of psychotherapy and sertraline treatment[40,41]. For this study, PTSD cases and controls were selected from the pre-treatment visit.

*Boston VA National Center for PTSD / Translational Research Center for TBI and Stress Disorders (NCPTSD/TRACTS)*. These analyses were performed using two cohorts comprising samples from three studies collected at the Boston VA Healthcare System. The VA Boston National Center for PTSD (NCPTSD) cohort includes subjects from two studies, the NCPTSD study (Miller PI)[42], and the PTSD & Accelerated Aging study (Wolf PI, Wolf et al., under review[43]). The NCPTSD study included trauma-exposed veterans and a subset of their trauma-exposed intimate partners. The PTSD & Accelerated Aging study included Veterans who screened positive for PTSD and who were subsequently evaluated comprehensively. The Translational Research Center for TBI and Stress Disorders (TRACTS)-study cohort (Milberg and Fortier PIs)[44] includes post-9/11 Veterans recruited for a study of TBI. For all three studies, PTSD diagnostic status was determined based on the Clinician-Administered PTSD Scale (CAPS) for DSM-IV[10] or DSM-5[45] and the Traumatic Life Events Questionnaire (TLEQ)[46] was used to assess adult and childhood trauma. The NCPTSD and TRACTS cohort DNAm data were jointly cleaned and analyzed together. For both cohorts, DNAm was measured at the Central Arkansas Veterans Healthcare System Pharmacogenomics Analysis Laboratory (PAL) using the Illumina Infinium MethylationEPIC BeadChip according to the manufacturer's protocols. Data cleaning and analyses were performed using the PGC-PTSD Workgroup quality control/analysis pipeline (https://github.com/PGC-PTSD-EWAS/EPIC_QC).

*Injury and Traumatic Stress (INTRuST)*. Patients and healthy controls were recruited from INTRuST clinical trials (with DNA collected prior to treatment) or specifically to contribute to the INTRuST biorepository. Patients were diagnosed with PTSD using study-specific measures which could have included either the CAPS-IV or CAPS-5, the MINI, or self-report using the PCL. Healthy controls were determined to be free of major psychiatric diagnoses, including PTSD, using the MINI or a comparable clinical interview. The Human Research Protection Program (HRPP) at UCSD approved this study, as did all the IRBs at participating sites.

*Mid-Atlantic Mental Illness Research Education and Clinical Center PTSD Study (VA-M-AA & VA-M-EA)*. As described previously[47], PTSD was diagnosed using the Structured Clinical Interview for DSM-IV Disorders (SCID) administered by trained interviewers. In accordance with the DSM-IV, PTSD consists of three symptom clusters. These include re-experiencing symptoms (B symptoms), avoidance and numbing symptoms (C symptoms) and hyperarousal symptoms (C symptoms). Total PTSD symptoms and symptom clusters (B, C, or D) were measured using the Davidson Trauma Scale for all veterans including individuals with current PTSD diagnosis and controls. The research was reviewed and approved by the Institutional Review Boards at the Salisbury VA, Hampton VA, Durham VA and Duke University Medical Centers.

**DNA methylation Quality Control of studies that used 450K array**

The QC of 9 studies that used 450K array is previously described in the study of Smith et al.[48] and available on https://github.com/PGC-PTSD-EWAS/QC_Pipeline. Briefly, ß-values, representative of the proportion of methylation at each probe, were calculated for all CpG sites in each sample. Samples with probe detection call rates <90% and those with an average intensity value of either <50% of the experiment-wide sample mean or <2000 arbitrary units (AU) were excluded. Probes with detection p-values > 0.001 or those based on less than three beads were set to missing as were probes that cross-hybridized between autosomes and sex chromosomes[49]. CpG sites with missing data for >10% of samples within cohorts were excluded from analysis. Normalization of probe distribution was conducted using Beta Mixture Quantile Normalization (BMIQ)[49] after background correction. Density plots of Type I and Type II probes before and after normalization were examined to confirm probe distributions between types were similar. ComBat was used to account for sources of technical variation including batch and positional effects, while preserving variation attributable to study-specific outcomes and covariates that would be used in downstream analyses (e.g., case status or sex). Proportions of CD8, CD4, NK, B cells, monocytes, and granulocytes were estimated using each individual's DNA methylation data based on the approach described by Houseman and colleagues and publicly available reference data (GSE36069)[50,51].

**Statistical analysis.** Within each cohort, logit transformed ß-values (M-values) were modeled by linear regression as a function of PTSD, adjusting for sex (if applicable), age, CD8, CD4, NK, B cell, and monocyte cell proportions, and ancestry using principal components (PCs). For cohorts with available GWAS data (Army STARRS, GTP, INTRuST, MRS, VA-M, VA-NCPTSD), PCs 1-3 were included as covariates. For cohorts without GWAS data (DNHS, PRISMO, WTC), the method described by Barfield and colleagues was used to generate ancestry PCs directly from the methylation data using CpG sites from the HumanMethylation450 array that are located within 0–100 bp of 1000 Genomes Project variants with minor allele frequency > 0.01, and PCs 2–4, which were the components that correlate most with ancestry, were used to adjust for ancestry as recommended by the authors[52]. Using the empirical Bayes method in the R package limma[53], moderated t-statistics were calculated for each CpG site, converted first into one-sided p-values then converted into z-scores to account for direction of effects.

**Ancestry Principal Components**

Ancestry principal components (PCs) were generated directly from DNA methylation data, using CpG sites from the HumanMethylation450 array that are located directly on 1000 Genomes Project variants with minor allele frequency >0.01, following the method described by Barfield and colleagues[52] as previously implemented[54]. Consistent with previous reports, PC1 was associated with other factors (e.g., cell type composition, smoking)[52,55].

**Smoking Score**

We used the weights of an EWAS of smoking[56] to generate smoking score from DNA methylation data. The weights include effect sizes of 39 CpGs that were associated with smoking pack years. We computed smoking score by calculating product of the M values for these 39 CpG sites times the effect size estimates.

**Postmortem brain DNAm**

*National center for PTSD Brain bank (NCPTSD-BB)*. We examined the associations with PTSD in dlPFC and vmPFC of 42 PTSD cases and 30 controls from the NCPTSD-BB using linear regressions, adjusting for age at death, sex, 3 ancestry PCs and the % neurons as estimated using the CETS package[57].

*PsychENCODE Consortium for PTSD (PEC-PTSD) Brainomics*. The associations between DNAm and PTSD in amygdala and DG were tested in 77 PTSD cases and 77 psychopathology-free controls from the PEC-PTSD. Two batches of data were analyzed (batch 1 had 50 cases and 50 controls, while batch 2 had 27 cases and 27 controls). Funnorm (*minfi* package[58]) was used for normalization, *ENmix*[59] for neuronal and non-neuronal cell type proportion estimation, *limma* package[60] for differential methylation. *Metafor* package[61] was used for fixed-effect meta-analysis of the results from the two batches.

**RNAseq Data**

*BEAR.* RNA was isolated by MagMAX for Stabilized Blood Tubes RNA Isolation Kit. All sequenced RNA samples had RNA Integrity Number (RIN) > 7. Library preparation was performed using the Illumina TruSeq Stranded Total RNA With Illumina Ribo-ZeroTM Plus rRNA reduction. The libraries were sequenced using a NextSeq550, which generated paired-end 100 bp reads. STAR is used to perform alignment against the hg38 human reference genome and transcriptome quantification[62]. Quality of the raw, trimmed, and aligned reads were checked using FastQC (https://www.bioinformatics.babraham.ac.uk/projects/fastqc/) and MultiQC[63] separately. Outlier samples (samples that are 1.5 times the interquartile range less than the first quartile [Q1] or 1.5 times the interquartile range greater than the third quartile [Q3]) in terms of uniquely mapped read counts and percentage of uniquely mapped reads. Self-report sex was confirmed by examining chrX gene *XIST* and chrY genes *EIF1AY*, *RPS4Y1*, *DDX3Y*, and *KDM5D*. Reads were normalized by log2 transformation in DEseq2[64].

*MRS.* RNA sequencing methods for MRS is described previously[65]. Briefly, RNA was isolated from peripheral blood leukocytes using LeukoLOCK Total RNA isolation. All mRNA samples passed an RNA integrity number (RIN) > 7. mRNA was subjected to Poly-A enrichment and 50bp paired-end sequencing on the Illumina Hi-Seq 2000. RNA libraries were prepared for sequencing using standard Illumina Tru-Seq protocols. Sequenced reads were trimmed for adaptor sequence and masked for low-complexity or low quality sequence, then mapped to hg19 RefSeq Human Genome and counted using RSEM[66]. Genes with low read counts were filtered in an unspecific manner using edgeR[67]. Genes having > 20 counts per million in at least 50% of the samples were retained. Resulting count data was normalized using the edgeR VOOM function[67,68] and subjected to clustering analysis and principal component analysis (PCA). Clustering, based on Pearson correlation and average distance metric, was used to spot outliers if outside 2.5 standard deviations from the average. Outliers were removed if outside 2 standard deviations from the average.

*AURORA.* Blood was collected using PAXgene RNA tubes. Total RNA was isolated using PAXgene blood miRNA kits (Qiagen, Germantown, MD) and RNA was stored at -80 ˚C until use. RNA concentration and purity were measured using Qubit RNA Assay Kits (ThermoFisher, Waltham, MA) and a 4200 TapeStation System (Agilent Technologies, Santa Clara, CA). The average RNA integrity (RIN) score across the cohort was 7.9, with 92% of samples having a RIN score ≥7. Four samples had RIN values below 4 and were discarded while the remainder were used to prepare RNA sequencing libraries. Template libraries for total RNA sequencing were produced from 600ng total RNA using Genomics Universal Plus Total RNA Seq Library kits with rRNA and globin depletion (TECAN, Morrisville, NC) according to manufacturer's specifications. Libraries were multiplexed in groups of 96 and sequenced on an NovaSeq S4 200 cycle kit (Illumina, San Diego, CA) using paired end reads (100x) at the University of North Carolina at Chapel Hill High Throughput Sequencing Facility. RNA-seq reads were trimmed, mapped to the hg38 human reference genome using STAR2.7.6a[62], then quantified using Salmon 1.4.0[69]. Raw read counts were then upper-quantile normalized and log2 transformed. Only genes that had at least 10 reads in any given library were used in subsequent analysis. Outliers were identified via principal component analysis. To verify the appropriate male/female categorization based on genetic attributes, median *XIST* RNA read counts were compared between women and men.

*NCPTSD Merit.* The sample consists of 204 participants (52% current PTSD, 85% male, 69% white) with mean (SD) age 56.54 (13.14). RNA was extracted using the Qiagen Paxgene RNA kit. Samples containing sufficient RNA were proceeded for sequencing. The Nanodrop RNA protocol was used to determine RNA concentrations. RNA Integrity (RIN) values for each sample were calculated using a Tapestation 4200 using RNA screen tapes (Agilent, 5067-5576) and buffer (Agilent, 5067-5577). 200 ng of total RNA and 1 ul of a 1:1000 dilution of ERCC RNA Spike-in mix 1 (ThermoFisher, 4456740) was used as input into Illumina TruSeq Stranded Total RNA with Ribo-Zero Globin kit (Illumina, 20020613) for library preparation. The libraries were indexed with IDT for Illumina – TruSeq RNA UD Indexes (Illumina, 20022371). Pooled libraries were sequenced on an Illumina NextSeq 550 sequencer using 150 cycle NextSeq 500/550 High Output Kits v2.5 (Illumina, 20024907) to produce 10-20 million 75-bp paired-end reads per sample. Raw reads were trimmed using Trimmomatic[70] to eliminate adapters and short or low-quality reads. Trimmed reads were aligned to hg38 human reference genome[71] via STAR[62]. Quality of the raw, trimmed, and aligned reads were checked using FastQC (https://www.bioinformatics.babraham.ac.uk/projects/fastqc/), RSeQC[72], and MultiQC[63] separately. Samples with less than 50% uniquely mapped reads were excluded. Aligned reads were then quantified using RSEM[66]. Self-report sex was confirmed by examining Y chromosome gene expressions. Raw counts were regularized log (rLog) transformed using DESeq2[64]. Principal Component (PC) of rLog expression values were generated, and outliers were identified and removed if being 6 SDs away from the mean for any of the first 10 PCs. Genotypes were called from the aligned reads using the GATK HaplotypeCaller (https://gatk.broadinstitute.org/hc/en-us/articles/360037225632-HaplotypeCaller) and compared with chip-based genotypes to confirm sample concordance.

**eFigure 1. Forest Plots of PTSD-associated CpGs in each cohort.** The x-axis is the effect size for each association and includes the 95% confidence interval. Error bars represent the 95% confidence interval, and the center of the error bars represent effect sizes for the CpGs in each cohort. Filled circles indicates a nominally significant association ($p < 0.05$). Hollow circles indicate the lack of association ($p > 0.05$). The vertical line indicates an effect size of 0. Primary meta-analysis is shown in blue and sensitivity analysis for smoking is shown in red.

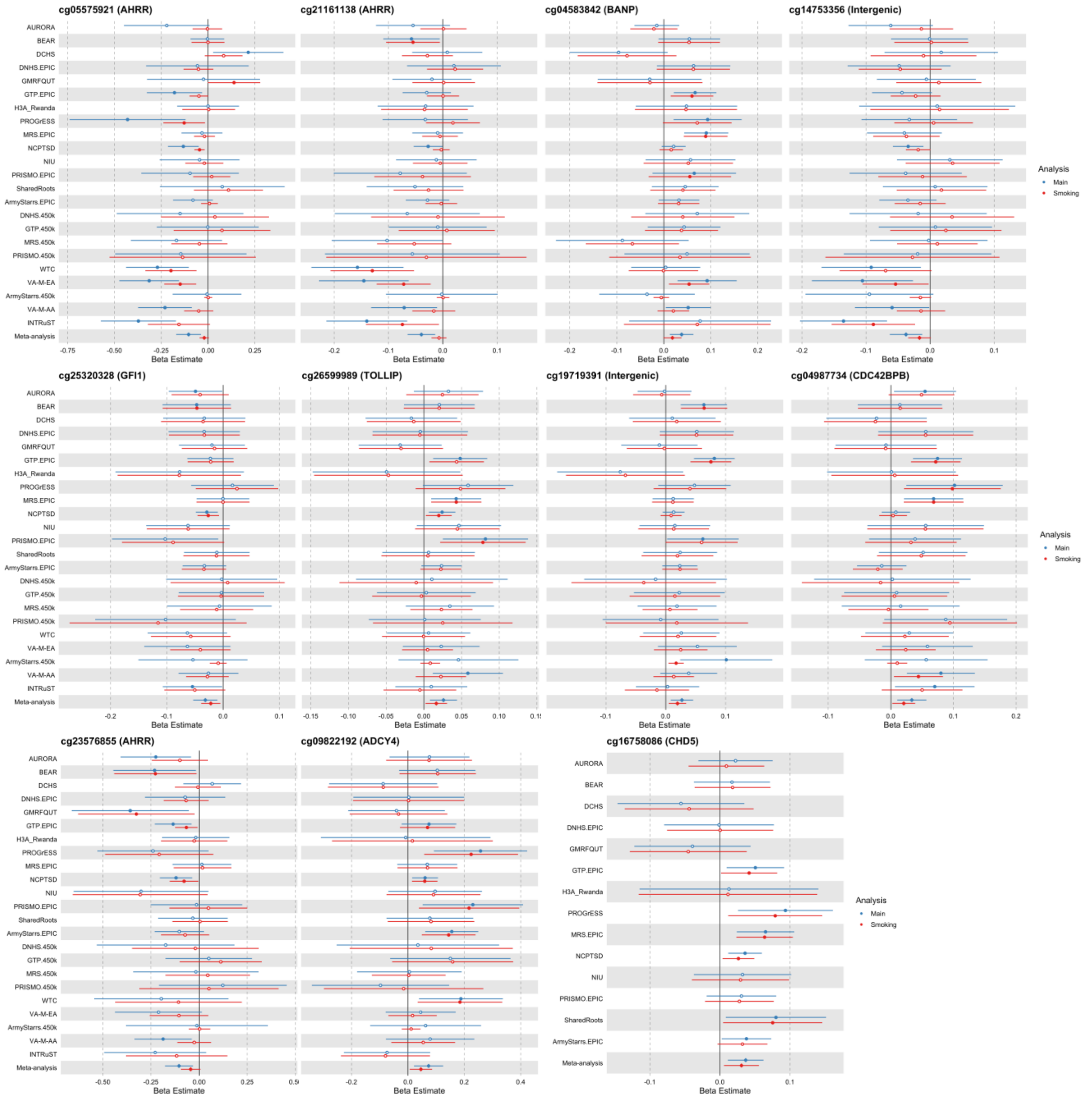

**eFigure 2. Manhattan plots of the stratified analyses.** Stratified analysis for **A)** females, **B)** males, **C)** European ancestry, **D)** African ancestry, **E)** civilian trauma, and **F)** military trauma. The x-axis depicts chromosomes and the location of each CpG site across the genome. The y-axis depicts the -log10 of the unadjusted p-value for the association with current PTSD. Each dot represents a CpG site. The solid blue line indicates the epigenome-wide statistical significance at p < 9e-8.

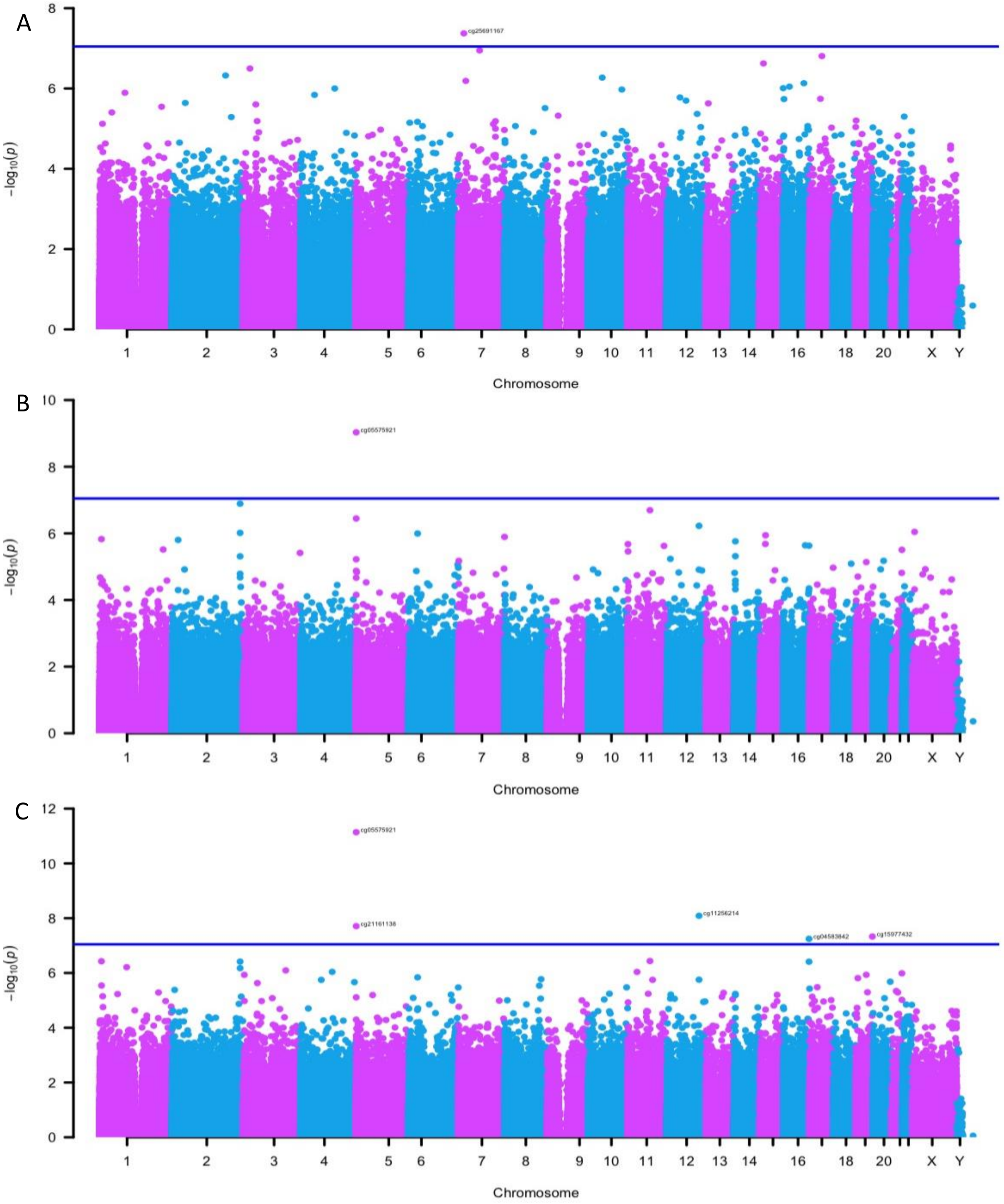

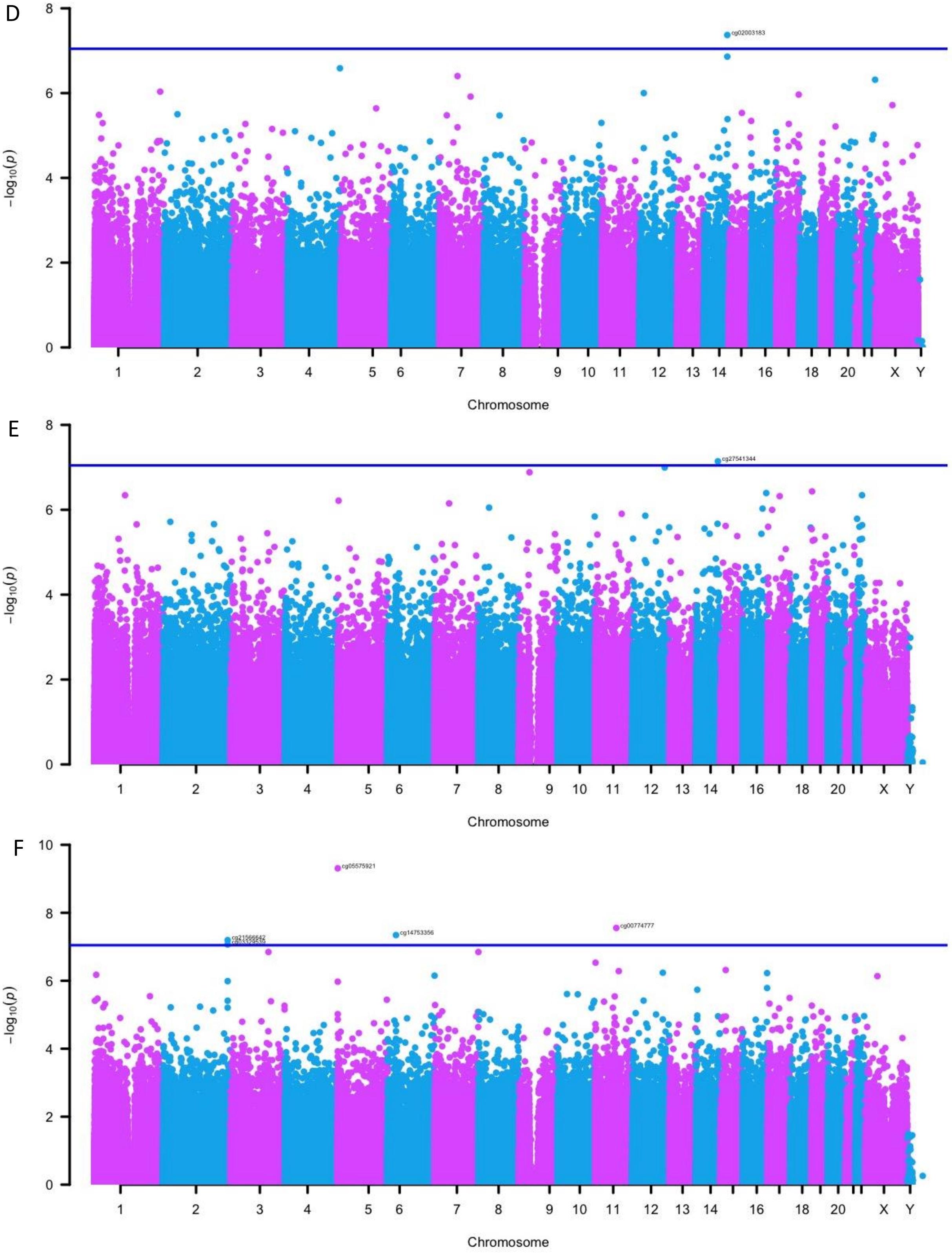
D
8
6
4
2
0
$-\log_{10}(p)$
cg02003183
1
2
3
4
5
6
7
8
9
10
11
12
13
14
16
18
20
X
Y
Chromosome
E
8
6
4
2
0
$-\log_{10}(p)$
cg27541344
1
2
3
4
5
6
7
8
9
10
11
12
13
14
16
18
20
X
Y
Chromosome
F
10
8
6
4
2
0
$-\log_{10}(p)$
cg05575921
cg21566642
cg14753356
cg00774777
1
2
3
4
5
6
7
8
9
10
11
12
13
14
16
18
20
X
Y
Chromosome

**eFigure 3. Forest Plots of CpGs associated with PTSD in the stratified analysis for females.** The x-axis is the effect size for each association and includes the 95% confidence interval. Error bars represent the 95% confidence interval, and the center of the error bars represent effect sizes for the CpGs in each cohort. Filled circles indicates a nominally significant association ($p < 0.05$). Hollow circles indicate the lack of association ($p > 0.05$). The vertical line indicates an effect size of 0.

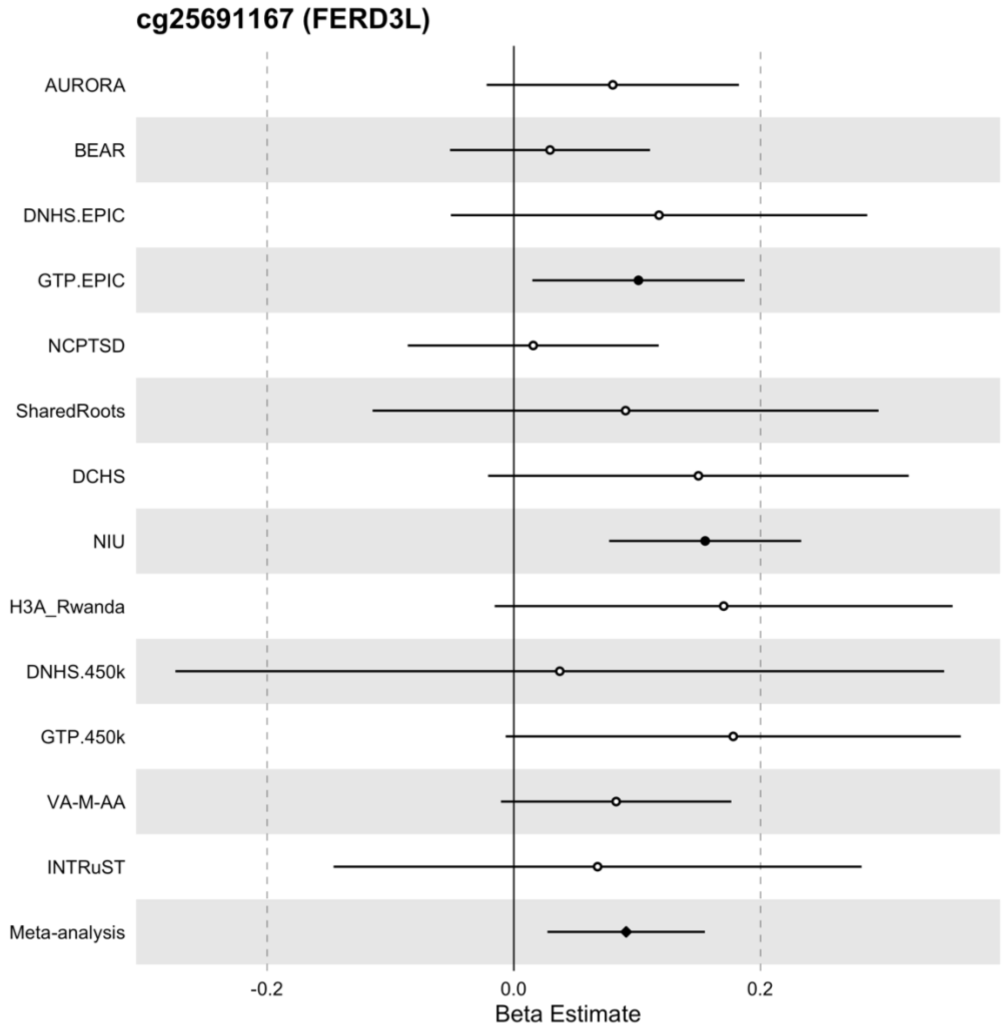

**eFigure 4. Forest Plots of CpGs associated with PTSD in the stratified analysis for males.** The x-axis is the effect size for each association and includes the 95% confidence interval. Error bars represent the 95% confidence interval, and the center of the error bars represent effect sizes for the CpGs in each cohort. Filled circles indicates a nominally significant association ($p < 0.05$). Hollow circles indicate the lack of association ($p > 0.05$). The vertical line indicates an effect size of 0.

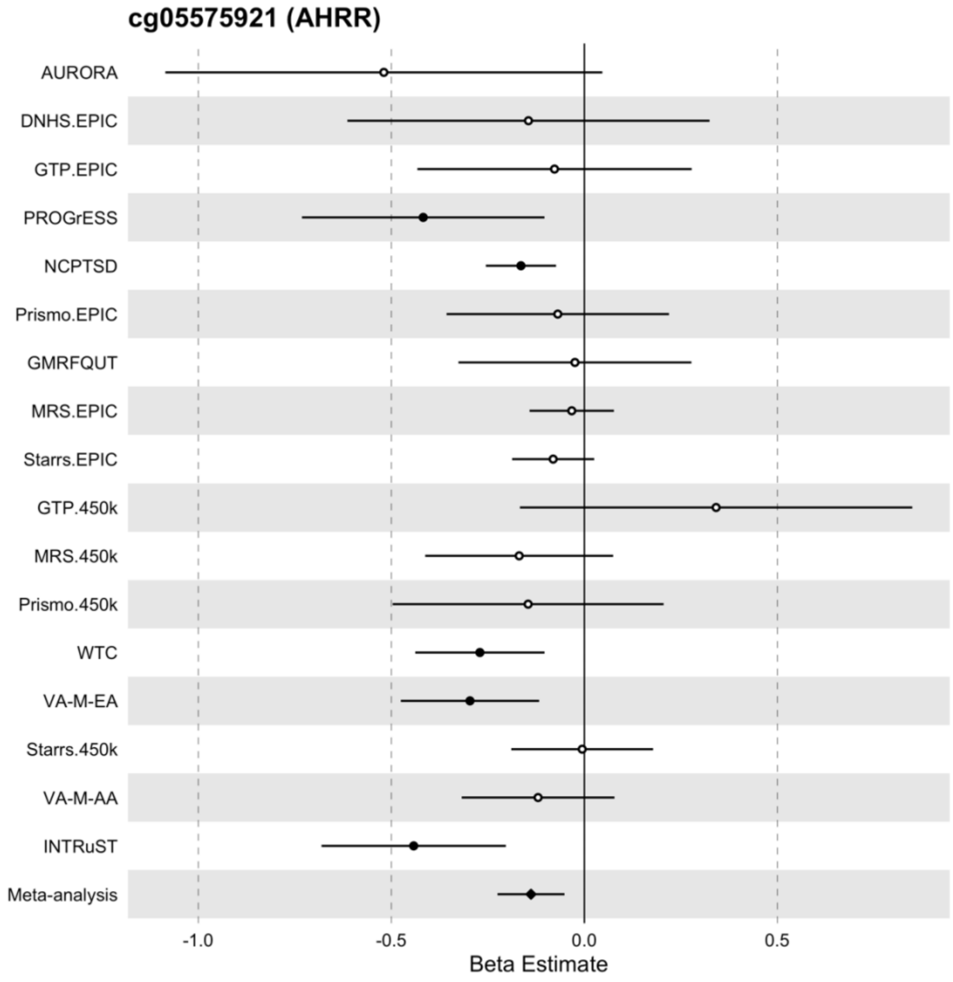

**eFigure 5. Forest Plots of CpGs associated with PTSD in the stratified analysis for European ancestry.** The x-axis is the effect size for each association and includes the 95% confidence interval. Error bars represent the 95% confidence interval, and the center of the error bars represent effect sizes for the CpGs in each cohort. Filled circles indicates a nominally significant association ($p < 0.05$). Hollow circles indicate the lack of association ($p > 0.05$). The vertical line indicates an effect size of 0.

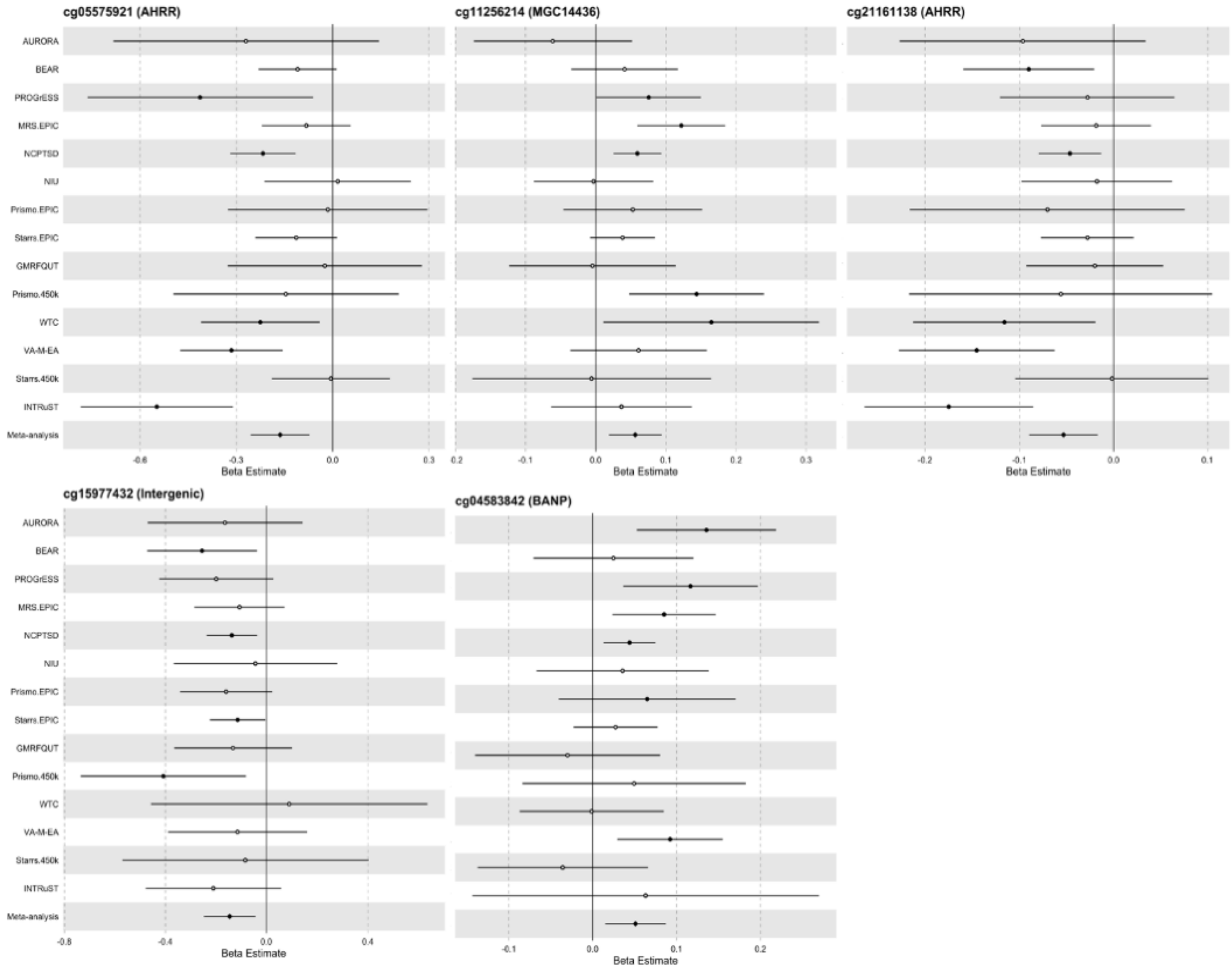

**eFigure 6. Forest Plots of CpGs associated with PTSD in the stratified analysis for African ancestry.** The x-axis is the effect size for each association and includes the 95% confidence interval. Error bars represent the 95% confidence interval, and the center of the error bars represent effect sizes for the CpGs in each cohort. Filled circles indicates a nominally significant association ($p < 0.05$). Hollow circles indicate the lack of association ($p > 0.05$). The vertical line indicates an effect size of 0.

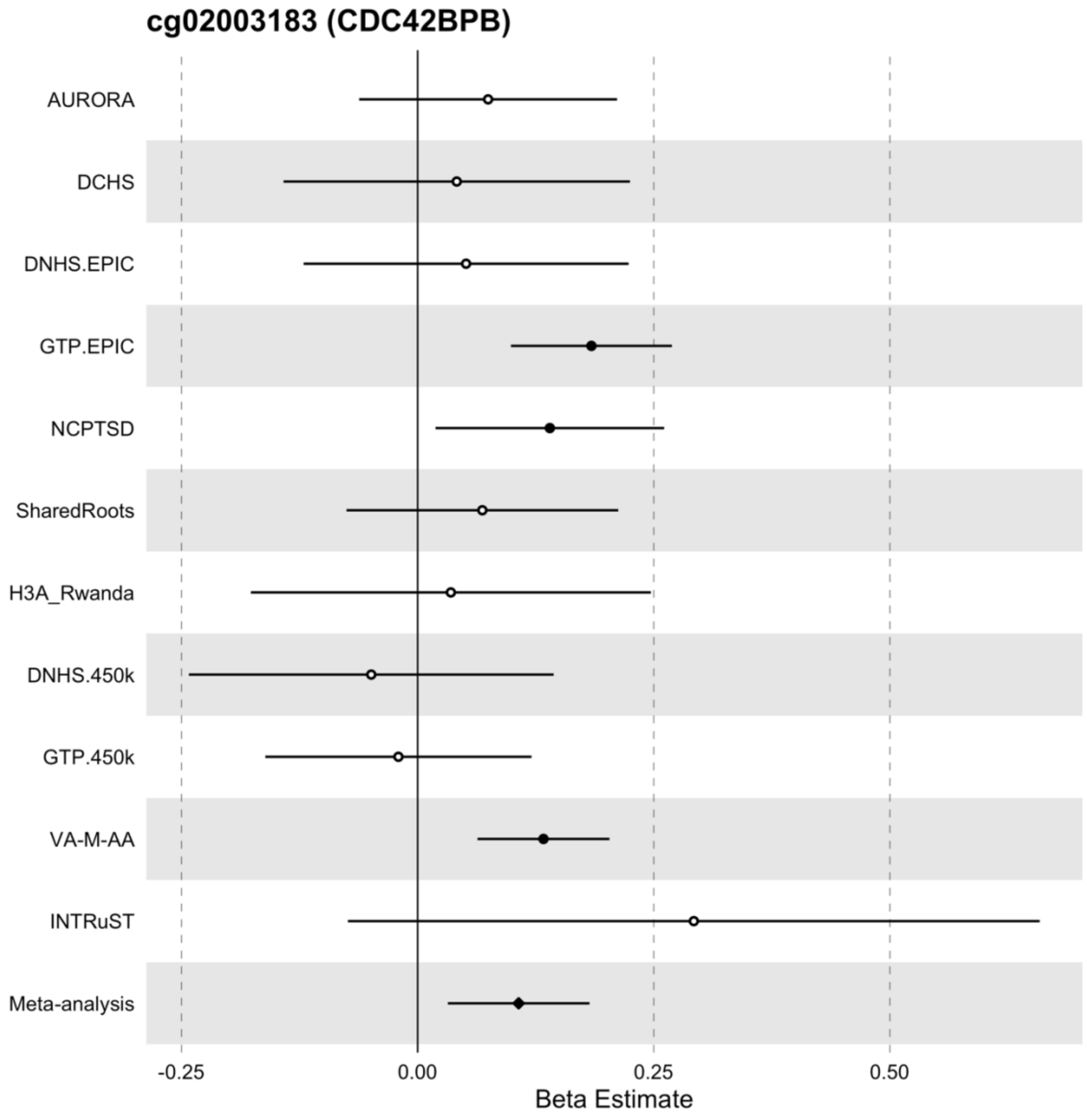

**eFigure 7. Forest Plots of CpGs associated with PTSD in the stratified analysis for civilian trauma.** The x-axis is the effect size for each association and includes the 95% confidence interval. Error bars represent the 95% confidence interval, and the center of the error bars represent effect sizes for the CpGs in each cohort. Filled circles indicates a nominally significant association ($p < 0.05$). Hollow circles indicate the lack of association ($p > 0.05$). The vertical line indicates an effect size of 0.

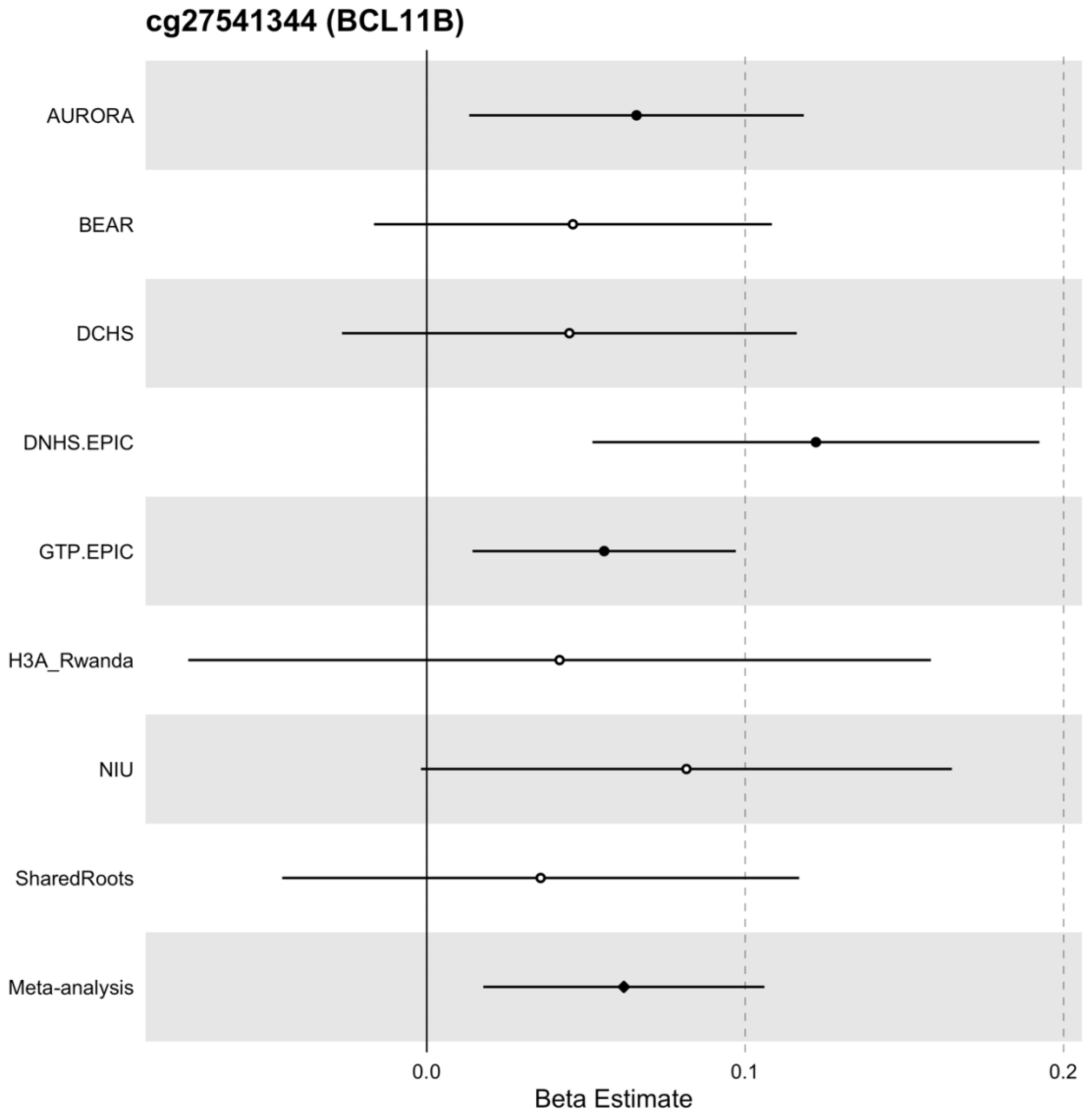

**eFigure 8. Forest Plots of CpGs associated with PTSD in the stratified analysis for military trauma.** The x-axis is the effect size for each association and includes the 95% confidence interval. Error bars represent the 95% confidence interval, and the center of the error bars represent effect sizes for the CpGs in each cohort. Filled circles indicates a nominally significant association ($p < 0.05$). Hollow circles indicate the lack of association ($p > 0.05$). The vertical line indicates an effect size of 0.

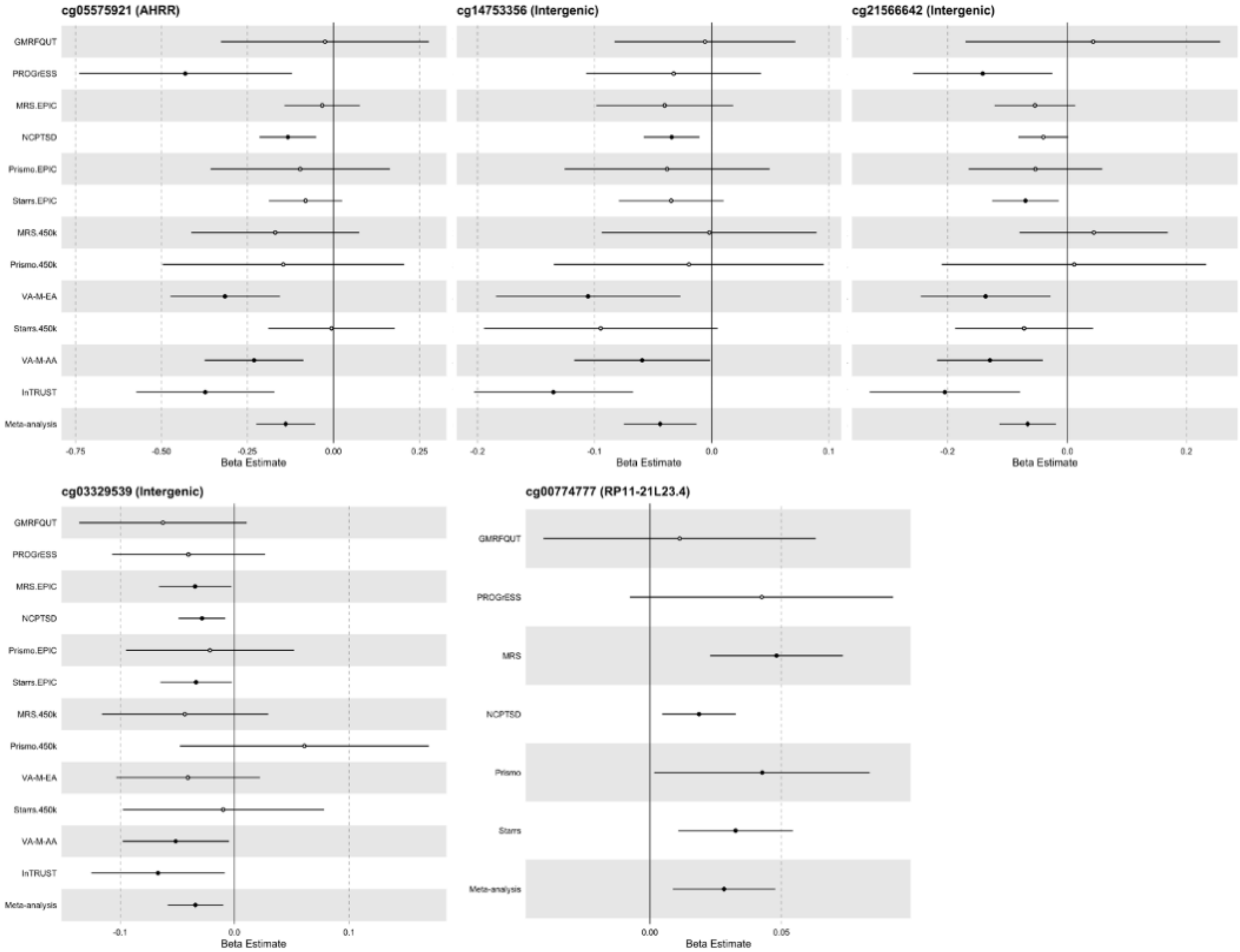

**eTable 1.** PTSD-associated CpGs from the primary analysis in the sensitivity analysis adjusted for smoking score.

| | | | Primary Analysis | | Sensitivity Analysis | |
|---|---|---|---|---|---|---|
| CpG | Position | Gene | z | *p*-value | z | *p*-value |
| cg05575921 | chr5:373378 | *AHRR* | -6.05 | 1.44E-09 | -3.17 | 1.52E-03 |
| cg21161138 | chr5:399360 | *AHRR* | -5.96 | 2.50E-09 | -1.94 | 0.053* |
| cg04583842 | chr16:88103117 | *BANP* | 5.81 | 6.29E-09 | 3.66 | 2.51E-04 |
| cg14753356 | chr6:30720108 | *Intergenic* | -5.72 | 1.09E-08 | -3.62 | 2.98E-04 |
| cg25320328 | chr1:92953037 | *GFI1* | -5.65 | 1.64E-08 | -4.96 | 7.10E-07 |
| cg26599989 | chr11:1297087 | *TOLLIP* | 5.58 | 2.35E-08 | 4.45 | 8.58E-06 |
| cg19719391 | chr4:26789915 | *Intergenic* | 5.58 | 2.45E-08 | 5.19 | 2.10E-07 |
| cg16758086 | chr1: 6173356 | *CHD5* | 5.55 | 2.85e-08 | 4.74 | 2.15E-06 |
| cg04987734 | chr14:103415873 | *CDC42BPB* | 5.53 | 3.26E-08 | 4.35 | 1.37E-05 |
| cg23576855 | chr5:373299 | *AHRR* | -5.52 | 3.44E-08 | -3.37 | 7.59E-04 |
| cg09822192 | chr14:24801191 | *ADCY4* | 5.44 | 5.30E-08 | 4.45 | 8.70E-06 |

Position is based on hg19. The sites that were not nominally significant ($p > 0.05$) in the sensitivity analysis adjusted for smoking score were indicated by an asterisk (*).

**eTable 2.** PTSD-associated CpGs from the primary analysis in the stratified analysis for sex, ancestry, and trauma type.

| | | **Primary Analysis** | | **Males** | | **Females** | | **European** | | **African** | | **Civilian** | | **Military** | |
|---|---|---|---|---|---|---|---|---|---|---|---|---|---|---|---|
| IlmnID | Gene | **z** | ***p*-value** | **z** | ***p*-value** | **z** | ***p*-value** | **z** | ***p*-value** | **z** | **p-value** | **z** | ***p*-value** | **z** | ***p*-value** |
| cg05575921 | *AHRR* | -6.05 | **1.44E-09** | -6.12 | **9.30E-10** | -2.21 | 2.72E-02 | -6.85 | **7.24E-12** | -1.22 | 2.21E-01* | -1.79 | 7.42E-02* | -6.22 | **4.92E-10** |
| cg21161138 | *AHRR* | -5.96 | **2.50E-09** | -5.09 | 3.59E-07 | -3.52 | 4.35E-04 | -5.62 | **1.95E-08** | -1.65 | 9.85E-02* | -3.26 | 1.11E-03 | -4.88 | 1.07E-06 |
| cg04583842 | *BANP* | 5.81 | **6.29E-09** | 4.08 | 4.50E-05 | 4.40 | 1.07E-05 | 5.43 | **5.66E-08** | 3.20 | 1.39E-03 | 3.23 | 1.22E-03 | 4.99 | 5.95E-07 |
| cg14753356 | *Intergenic* | -5.72 | **1.09E-08** | -4.89 | 1.01E-06 | -3.07 | 2.18E-03 | -4.82 | 1.45E-06 | -1.39 | 1.65E-01* | -1.88 | 6.04E-02* | -5.47 | **4.53E-08** |
| cg25320328 | *GFI1* | -5.65 | **1.64E-08** | -4.08 | 4.59E-05 | -3.82 | 1.35E-04 | -4.99 | 6.15E-07 | -2.08 | 3.71E-02 | -3.78 | 1.55E-04 | -4.37 | 1.24E-05 |
| cg26599989 | *TOLLIP* | 5.58 | **2.35E-08** | 4.64 | 3.49E-06 | 2.78 | 5.51E-03 | 4.38 | 1.19E-05 | 3.20 | 1.39E-03 | 2.14 | 3.27E-02 | 5.13 | 2.94E-07 |
| cg19719391 | *Intergenic* | 5.58 | **2.45E-08** | 3.71 | 2.06E-04 | 4.07 | 4.79E-05 | 4.27 | 1.98E-05 | 3.37 | 7.43E-04 | 4.54 | 5.53E-06 | 3.59 | 3.34E-04 |
| cg16758086 | *CHD5* | 5.55 | **2.85E-08** | 4.81 | 1.49E-06 | 2.78 | 5.44E-03 | 5.08 | 3.74E-07 | 2.40 | 1.65E-02 | 2.60 | 9.38E-03 | 4.97 | 6.68E-07 |
| cg04987734 | *CDC42BPB* | 5.53 | **3.26E-08** | 3.59 | 3.27E-04 | 3.74 | 1.83E-04 | 3.66 | 2.49E-04 | 5.27 | 1.38E-07 | 4.15 | 3.29E-05 | 3.81 | 1.39E-04 |
| cg23576855 | *AHRR* | -5.52 | **3.44E-08** | -4.53 | 6.01E-06 | -3.62 | 2.97E-04 | -4.05 | 5.06E-05 | -2.09 | 3.67E-02 | -3.27 | 1.07E-03 | -4.43 | 9.61E-06 |
| cg09822192 | *ADCY4* | 5.44 | **5.30E-08** | 4.78 | 1.73E-06 | 2.15 | 3.12E-02 | 4.53 | 5.81E-06 | 2.03 | 4.27E-02 | 3.59 | 3.28E-04 | 4.40 | 1.06E-05 |
| cg25691167 | *FERD3L* | 3.57 | 3.60E-04 | 0.57 | 5.67E-01* | 5.48 | **4.24E-08** | 2.51 | 1.21E-02 | 3.19 | 1.43E-03 | 4.05 | 5.12E-05 | 1.50 | 1.33E-01* |
| cg11256214 | *MGC14436* | 4.83 | 1.36E-06 | 4.99 | 5.95E-07 | 1.45 | 1.47E-01* | 5.76 | **8.17E-09** | 1.50 | 1.34E-01* | 1.17 | 2.42E-01* | 5.00 | 5.80E-07 |
| cg15977432 | *Intergenic* | -3.59 | 3.31E-04 | -3.51 | 4.56E-04 | -1.34 | 1.81E-01* | -5.46 | **4.68E-08** | -0.87 | 3.87E-01* | -0.49 | 6.27E-01* | -3.51 | 4.55E-04 |
| cg02003183 | *CDC42BPB* | 3.99 | 6.65E-05 | 2.22 | 2.63E-02 | 3.79 | 1.52E-04 | 2.33 | 1.98E-02 | 5.48 | **4.26E-08** | 3.19 | 1.43E-03 | 2.74 | 6.18E-03 |
| cg27541344 | *BCL11B* | 3.96 | 7.38E-05 | 2.13 | 3.33E-02 | 3.49 | 4.74E-04 | 1.43 | 1.54E-01* | 3.54 | 3.95E-04 | 5.39 | **7.21E-08** | 0.88 | 3.77E-01* |
| cg00774777 | *RP11-21L23.4* | 4.93 | 8.24E-07 | 5.20 | 2.02E-07 | 1.09 | 2.77E-01* | 5.09 | 3.67E-07 | 0.75 | 4.56E-01* | 0.71 | 4.76E-01* | 5.55 | **2.79E-08** |
| cg21566642 | *Intergenic* | -5.25 | 1.52E-07 | -5.28 | 1.29E-07 | -2.23 | 2.56E-02 | -5.08 | 3.85E-07 | -1.06 | 2.87E-01* | -1.31 | 1.89E-01* | -5.41 | **6.36E-08** |
| cg03329539 | *Intergenic* | -5.02 | 5.27E-07 | -4.90 | 9.70E-07 | -2.10 | 3.61E-02 | -4.06 | 4.83E-05 | -0.86 | 3.88E-01* | -1.01 | 3.12E-01* | -5.36 | **8.53E-08** |

Epigenome-wide significant sites ($p < 9e-08$) were shown in bold. Non-significant sites ($p > 0.05$) were indicated by an asterisk (*).

**eTable 3.** Gene Ontology Enrichment results in females.

| GO ID | GO term | N | DE | P.DE | FDR |
|---|---|---|---|---|---|
| GO:0007399 | nervous system development | 2474 | 175 | 7.34E-09 | 1.17E-04 |
| GO:0048731 | system development | 3971 | 238 | 5.27E-08 | 4.20E-04 |
| GO:0007275 | multicellular organism development | 4643 | 260 | 6.16E-07 | 3.27E-03 |

GO: Gene Ontology. N: Number of genes in the GO term. DE: Number of genes that are differentially methylated. P.DE: p-value for over-representation of the GO term. FDR: False discovery rate.

**eTable 4.** Correlation values of methylation between blood and brain regions from Blood Brain DNA Methylation Comparison Tool.

| | | PFC | | EC | | STG | | CER | |
|---|---|---|---|---|---|---|---|---|---|
| CpG | Gene | r | *p*-value | r | *p*-value | r | *p*-value | r | *p*-value |
| cg05575921 | *AHRR* | **0.280** | **0.016** | 0.194 | 0.104 | 0.208 | 0.0735 | -0.008 | 0.948 |
| cg21161138 | *AHRR* | 0.177 | 0.131 | 0.142 | 0.239 | **0.282** | **0.014** | -0.084 | 0.486 |
| cg04583842 | *BANP* | -0.181 | 0.122 | 0.126 | 0.297 | **0.390** | **5.48e-04** | 0.001 | 0.990 |
| cg14753356 | *Intergenic* | 0.050 | 0.673 | 0.066 | 0.583 | 0.102 | 0.382 | -0.044 | 0.713 |
| cg25320328 | *GFI1* | 0.198 | 0.091 | -0.148 | 0.217 | 0.009 | 0.939 | 0.009 | 0.938 |
| cg26599989 | *TOLLIP* | 0.0505 | 0.669 | -0.074 | 0.539 | 0.177 | 0.129 | 0.160 | 0.183 |
| cg19719391 | *Intergenic* | 0.148 | 0.208 | **0.256** | **0.031** | -0.001 | 0.992 | 0.102 | 0.397 |
| cg16758086 | *CHD5* | NA | NA | NA | NA | NA | NA | NA | NA |
| cg04987734 | *CDC42BPB* | -0.053 | 0.652 | 0.108 | 0.369 | 0.184 | 0.114 | -0.021 | 0.865 |
| cg23576855 | *AHRR* | **0.906** | **1.51e-28** | **0.86** | **6.85e-22** | **0.890** | **1.18e-26** | **0.867** | **1.39e-22** |
| cg09822192 | *ADCY4* | 0.142 | 0.226 | 0.179 | 0.136 | 0.053 | 0.653 | 0.223 | 0.062 |
| cg25691167 | *FERD3L* | -0.056 | 0.637 | 0.022 | 0.855 | -0.107 | 0.363 | -0.211 | 0.078 |
| cg11256214 | *MGC14436* | -0.058 | 0.623 | 0.052 | 0.670 | 0.040 | 0.731 | 0.033 | 0.786 |
| cg15977432 | *Intergenic* | **0.751** | **1.37e-14** | **0.799** | **6.43e-17** | **0.808** | **1.95e-18** | **0.754** | **3.32e-14** |
| cg02003183 | *CDC42BPB* | 0.130 | 0.268 | 0.192 | 0.108 | **0.263** | **0.023** | 0.144 | 0.232 |
| cg27541344 | *BCL11B* | NA | NA | NA | NA | NA | NA | NA | NA |
| cg00774777 | *RP11-21L23.4* | NA | NA | NA | NA | NA | NA | NA | NA |
| cg21566642 | *Intergenic* | **0.386** | **6.82e-04** | **0.384** | **9.39e-04** | **0.508** | **3.28e-06** | 0.117 | 0.329 |
| cg03329539 | *Intergenic* | -0.065 | 0.58 | -0.057 | 0.639 | **0.301** | **0.009** | 0.173 | 0.143 |

The list includes CpGs identified from the primary meta-analysis and the stratified analyses for sex, ancestry, and trauma type. PFC: prefrontal cortex. EC: entorhinal cortex. STG: superior temporal gyrus. CER: cerebellum. NA indicates that CpG is not available in the Blood Brain DNA Methylation Comparison Tool. Significant correlations are shown in bold.

**eTable 5.** Results of PTSD-associated CpGs from blood in the brain.

| | | dlPFC | | vmPFC | | Amygdala | | DG | |
|---|---|---|---|---|---|---|---|---|---|
| CpG | Gene | logFC | *p*-value | logFC | *p*-value | logFC | *p*-value | logFC | *p*-value |
| cg05575921 | *AHRR* | -0.03 | 0.725 | -0.06 | 0.451 | -0.01 | 0.922 | 0.16 | 0.127 |
| cg21161138 | *AHRR* | -0.24 | **5.92E-03** | 0.06 | 0.512 | 0.01 | 0.937 | -0.09 | 0.186 |
| cg04583842 | *BANP* | 0.08 | 0.466 | 0.04 | 0.612 | 0.04 | 0.630 | -0.06 | 0.495 |
| cg14753356 | *Intergenic* | 0.25* | **7.60E-03** | 0.25* | **0.016** | 0.15 | 0.110 | 0.04 | 0.638 |
| cg25320328 | *GFI1* | 0.18* | **0.036** | -0.06 | 0.453 | -0.03 | 0.544 | -0.02 | 0.787 |
| cg26599989 | *TOLLIP* | -0.06 | 0.427 | -0.03 | 0.625 | -0.08 | 0.077 | -0.02 | 0.631 |
| cg19719391 | *Intergenic* | 0.08 | 0.353 | 0.09 | 0.329 | -0.02 | 0.664 | 0.16 | **0.003** |
| cg16758086 | *CHD5* | 0.02 | 0.856 | 0.03 | 0.818 | 0.05 | 0.475 | -0.06 | 0.400 |
| cg04987734 | *CDC42BPB* | 0.20 | **3.90E-03** | 0.05 | 0.445 | 0.05 | 0.527 | 0.08 | 0.209 |
| cg23576855 | *AHRR* | -0.06 | 0.699 | -0.10 | 0.527 | NA | NA | NA | NA |
| cg09822192 | *ADCY4* | 0.02 | 0.746 | -0.04 | 0.581 | 0.04 | 0.332 | 0.00 | 0.952 |
| cg25691167 | *FERD3L* | -0.09 | 0.249 | -0.02 | 0.784 | -0.10 | 0.398 | 0.07 | 0.494 |
| cg11256214 | *MGC14436* | 0.02 | 0.812 | 0.12 | 0.054 | 0.01 | 0.748 | -0.05 | 0.119 |
| cg15977432 | *Intergenic* | -0.69 | 0.154 | -0.23 | 0.640 | NA | NA | NA | NA |
| cg02003183 | *CDC42BPB* | 0.17 | 0.192 | 0.07 | 0.557 | 0.16 | 0.060 | 0.07 | 0.551 |
| cg27541344 | *BCL11B* | 0.04 | 0.662 | -0.05 | 0.708 | 0.02 | 0.786 | -0.10 | 0.196 |
| cg00774777 | *RP11-21L23.4* | -0.16 | 0.152 | 0.01 | 0.938 | 0.15 | **0.039** | 0.33 | **3.79E-06** |
| cg21566642 | *Intergenic* | -0.13 | 0.112 | -0.09 | 0.274 | 0.00 | 0.991 | 0.21* | **6.86E-03** |
| cg03329539 | *Intergenic* | -0.07 | 0.294 | 0.15 | 0.082 | -0.11 | 0.055 | -0.08 | 0.191 |

The list includes CpGs identified from the primary meta-analysis and the stratified analyses for sex, ancestry, and trauma type. dlPFC: dorsolateral prefrontal cortex. vmPFC: ventromedial prefrontal cortex. DG: dentate gyrus. logFC: logarithm of fold change. NA indicates that CpG was not available in the post-mortem brain dataset. Significant associations with PTSD are shown in bold. Sites with an opposite direction of association in the brain were indicated with an asterisk (*).

**eTable 6.** Results of PTSD-associated CpGs from blood in neuronal nuclei.

| | | | 5mC | | | 5hmC | | |
|---|---|---|---|---|---|---|---|---|
| CpG | Position | Gene | Position | ΔDNAm | *p*-value | Position | ΔDNAm | *p*-value |
| cg05575921 | chr5:373263 | *AHRR* | chr5:373668 | -2.21 | 4.84E-04 | chr5:373710 | 5.29* | 2.57E-03 |
| | | | chr5:373673 | 1.37* | 9.11E-03 | | | |
| cg21161138 | chr5:399245 | *AHRR* | chr5:399720 | -3.40 | 0.045 | | | |
| cg25691167 | chr7:19145338 | *FERD3L* | chr7:19145258 | 0.67 | 3.25E-03 | | | |
| cg21566642 | chr2:232419951 | *Intergenic* | chr2:232419004 | 0.47* | 0.026 | chr2:232418992 | 0.37* | 0.045 |
| | | | chr2:232419552 | -2.07 | 4.17E-03 | chr2:232419019 | 0.67* | 0.014 |
| | | | chr2:232419868 | -2.37 | 0.022 | chr2:232419555 | -1.97 | 0.026 |
| | | | | | | chr2:232419980 | 5.96* | 0.016 |
| | | | | | | chr2:232419981 | -5.56 | 0.047 |

Only significant associations were shown. Neuronal nuclei 5mC and 5hmC results span 500 bp upstream and downstream of PTSD-associated CpGs from blood. 5mC: 5-Methylcytosine. 5hmC: 5-Hydroxymethylcytosine. ΔDNAm: Mean DNA methylation difference between PTSD cases and controls. Position is based on hg38. Sites with an opposite direction of association in neuronal nuclei were indicated with an asterisk (*).

**eTable 7.** Results of PTSD-associated CpGs from blood in fibroblasts.

| CpG | Gene | ΔDNAm | t | *p*-value |
|---|---|---|---|---|
| cg05575921 | *AHRR* | -0.005 | -0.67 | 0.52 |
| cg21161138 | *AHRR* | 0.225* | 15.55 | **5.76E-07** |
| cg04583842 | *BANP* | -0.003 | -0.44 | 0.68 |
| cg14753356 | *Intergenic* | 0.012 | 0.81 | 0.44 |
| cg25320328 | *GFI1* | 0.024* | 2.42 | **0.04** |
| cg26599989 | *TOLLIP* | 0.067 | 6.40 | **1.31E-04** |
| cg19719391 | *Intergenic* | 0.057 | 5.27 | **4.01E-04** |
| cg16758086 | *CHD5* | 0.101 | 6.67 | **7.11E-04** |
| cg04987734 | *CDC42BPB* | 0.015 | 1.65 | 0.14 |
| cg23576855 | *AHRR* | NA | NA | NA |
| cg09822192 | *ADCY4* | 0.025 | 0.93 | 0.39 |
| cg25691167 | *FERD3L* | 0.104 | 5.26 | **6.53E-04** |
| cg11256214 | *MGC14436* | 0.114 | 6.94 | **5.76E-04** |
| cg15977432 | *Intergenic* | NA | NA | NA |
| cg02003183 | *CDC42BPB* | -0.010 | -0.53 | 0.61 |
| cg27541344 | *BCL11B* | -0.003 | -0.11 | 0.91 |
| cg00774777 | *RP11-21L23.4* | 0.005 | 0.94 | 0.37 |
| cg21566642 | *Intergenic* | -0.001 | -0.05 | 0.96 |
| cg03329539 | *Intergenic* | -0.034 | -1.69 | 0.15 |

The list includes CpGs identified from the primary meta-analysis and the stratified analyses for sex, ancestry, and trauma type. ΔDNAm: Mean DNA methylation difference between glucocorticoid treatment and vehicle. NA indicates that CpG was not available in the fibroblast dataset. Significant associations with PTSD are shown in bold. Sites with an opposite direction of association in the fibroblasts were indicated with an asterisk (*).

**eTable 8.** Blood DNA methylation and cis-gene expression (RNAseq) correlations.

| | | BEAR | | AURORA | | MRS | | NCPTSD Merit | | Meta-Analysis | |
|---|---|---|---|---|---|---|---|---|---|---|---|
| CpG | Gene | r | *p*-value | r | *p*-value | r | *p*-value | r | *p*-value | z | *p*-value |
| cg05575921 | *AHRR* | -0.18 | **0.04** | -0.67 | **<2.20E-16** | -0.6 | **3.25E-36** | -0.38 | **2.71E-08** | -16.69 | **1.67E-62** |
| cg21161138 | *AHRR* | -0.02 | 0.80 | -0.59 | **<2.20E-16** | -0.48 | **4.04E-21** | -0.29 | **2.65E-05** | -12.45 | **1.36E-35** |
| cg04583842 | *BANP* | 0.22 | **0.01** | -0.04 | 0.59 | 0.2 | **1.65E-04** | -0.12 | 0.10 | 2.35 | **1.88E-02** |
| cg14753356 | *Intergenic* | NA | NA | NA | NA | NA | NA | NA | NA | NA | NA |
| cg25320328 | *GFI1* | 0.09 | 0.30 | -0.14 | 0.07 | 0.16 | **2.27E-03** | -0.12 | 0.08 | 0.69 | 0.49 |
| cg26599989 | *TOLLIP* | -0.06 | 0.52 | -0.21 | **0.007** | -0.07 | 0.21 | 0.03 | 0.64 | -2.14 | **3.20E-02** |
| cg19719391 | *Intergenic* | NA | NA | NA | NA | NA | NA | NA | NA | NA | NA |
| cg16758086 | *CHD5* | -0.17 | **0.05** | -0.01 | 0.9 | NA | NA | -0.02 | 0.73 | -1.21 | 0.22 |
| cg04987734 | *CDC42BPB* | 0.20 | **0.02** | 0.08 | 0.28 | -0.08 | 0.13 | -0.05 | 0.48 | 0.01 | 0.99 |
| cg23576855 | *AHRR* | 0.01 | 0.94 | NA | NA | -0.23 | **1.73E-05** | -0.22 | **1.74E-03** | -4.86 | **1.18E-06** |
| cg18846074 | *ADCY4* | 0.13 | 0.14 | 0.05 | 0.52 | -0.15 | **5.63E-03** | -0.07 | 0.35 | -1.47 | 0.14 |
| cg25691167 | *FERD3L* | NA | NA | NA | NA | NA | NA | NA | NA | NA | NA |
| cg11256214 | *MGC14436* | NA | NA | NA | NA | NA | NA | NA | NA | NA | NA |
| cg15977432 | *Intergenic* | NA | NA | NA | NA | NA | NA | NA | NA | NA | NA |
| cg02003183 | *CDC42BPB* | 0.10 | 0.27 | 0.12 | 0.12 | -0.09 | 0.10 | -0.12 | 0.10 | -0.80 | 0.43 |
| cg27541344 | *BCL11B* | -0.56 | **5.41E-12** | -0.52 | **3.49E-13** | NA | NA | **-0.50** | **1.54E-14** | -12.89 | **4.90E-38** |
| cg00774777 | *RP11-21L23.4* | NA | NA | NA | NA | NA | NA | NA | NA | NA | NA |
| cg21566642 | *Intergenic* | NA | NA | NA | NA | NA | NA | NA | NA | NA | NA |
| cg03329539 | *Intergenic* | NA | NA | NA | NA | NA | NA | NA | NA | NA | NA |

The list includes CpGs identified from the primary meta-analysis and the stratified analyses for sex, ancestry, and trauma type. NA indicates that CpGs or genes were not available in the datasets. Significant correlations are shown in bold.

**eTable 9.** Significant SNPs from cis-meQTL analysis and their association with PTSD.

| | | | | | | meQTL Analysis | | | | | GWAS | | | |
|---|---|---|---|---|---|---|---|---|---|---|---|---|---|---|
| **CpG** | **CpG Location** | **Gene** | **SNP** | **MAF** | **SNP Location** | **A1** | **Beta** | **SE** | ***p*-value** | **Source** | **A1** | **A2** | **z** | ***p*-value** |
| cg05575921 | chr5:373378 | *AHRR* | rs35947614 | 0.06 | chr5:224397 | G | 0.23 | 0.01 | <5E-324 | GoDMC | A | G | 0.27 | 0.79 |
| cg05575921 | chr5:373378 | *AHRR* | rs11958189 | 0.11 | chr5:398696 | A | 0.25 | 0.05 | 7.23E-08 | meQTL EPIC | A | G | -0.87 | 0.39 |
| cg21161138 | chr5:399360 | *AHRR* | rs2434697 | 0.21 | chr5:466811 | T | -0.16 | 0.01 | <5E-324 | GoDMC | T | C | -1.68 | 0.09 |
| cg04583842 | chr16:88103117 | *BANP* | rs13331671 | 0.15 | chr16:88009179 | A | -0.13 | 0.02 | 3.26E-13 | GoDMC | NA | NA | NA | NA |
| cg04583842 | chr16:88103117 | *BANP* | rs8050099 | 0.26 | chr16:87319096 | T | -0.35 | 0.07 | 1.11E-06 | meQTL EPIC | T | G | 1.01 | 0.31 |
| cg04583842 | chr16:88103117 | *BANP* | rs7194688 | 0.25 | chr16:87322169 | C | -0.32 | 0.07 | 1.60E-05 | meQTL EPIC | T | C | -1.21 | 0.23 |
| cg14753356 | chr6:30720108 | *Intergenic* | rs28986310 | 0.05 | chr6:29735797 | T | 0.31 | 0.02 | <5E-324 | GoDMC | T | C | 6.73 | **1.68E-11** |
| cg14753356 | chr6:30720108 | *Intergenic* | rs562745472 | 0.05 | chr6:30774269 | G | 0.26 | 0.03 | <5E-324 | GoDMC | NA | NA | NA | NA |
| cg14753356 | chr6:30720108 | *Intergenic* | rs2524212 | 0.38 | chr6:30345871 | C | -0.16 | 0.03 | 1.64E-06 | meQTL EPIC | NA | NA | NA | NA |
| cg14753356 | chr6:30720108 | *Intergenic* | rs28780104 | 0.08 | chr6:30424655 | G | 0.46 | 0.06 | 2.34E-14 | meQTL EPIC | C | G | -5.33 | **9.96E-08** |
| cg14753356 | chr6:30720108 | *Intergenic* | rs13214831 | 0.26 | chr6:30731505 | A | 0.49 | 0.04 | 4.08E-33 | meQTL EPIC | A | G | 4.96 | **7.23E-07** |
| cg14753356 | chr6:30720108 | *Intergenic* | rs377098668 | <0.001 | chr6:30748618 | delA | 0.34 | 0.04 | 2.32E-21 | meQTL EPIC | NA | NA | NA | NA |
| cg14753356 | chr6:30720108 | *Intergenic* | rs3131063 | 0.44 | chr6:30763756 | A | -0.20 | 0.04 | 1.26E-08 | meQTL EPIC | A | G | -2.27 | **2.33E-02** |
| cg14753356 | chr6:30720108 | *Intergenic* | rs12206683 | 0.42 | chr6:30771086 | C | 0.17 | 0.03 | 1.38E-06 | meQTL EPIC | T | C | -2.98 | **2.92E-03** |
| cg14753356 | chr6:30720108 | *Intergenic* | rs9468851 | 0.50 | chr6:30930633 | G | 0.15 | 0.04 | 5.40E-05 | meQTL EPIC | NA | NA | NA | NA |
| cg14753356 | chr6:30720108 | *Intergenic* | rs2077492 | 0.50 | chr6:31606392 | A | 0.12 | 0.03 | 8.79E-05 | meQTL EPIC | A | G | 3.78 | **1.60E-04** |
| cg25320328 | chr1:92953037 | *GFI1* | rs11809700 | 0.27 | chr1:93152635 | T | -0.09 | 0.01 | <5E-324 | GoDMC | T | C | -0.83 | 0.40 |
| cg26599989 | chr11:1297087 | *TOLLIP* | rs56139112 | 0.05 | chr11:1257886 | T | -1.12 | 0.03 | <5E-324 | GoDMC | T | C | -1.17 | 0.24 |
| cg26599989 | chr11:1297087 | *TOLLIP* | rs55902810 | 0.06 | chr11:1248723 | T | -0.55 | 0.07 | 7.91E-16 | meQTL EPIC | T | C | -0.50 | 0.62 |
| cg26599989 | chr11:1297087 | *TOLLIP* | rs5743987 | 0.08 | chr11:1305846 | T | -0.70 | 0.06 | 2.49E-32 | meQTL EPIC | T | C | -0.54 | 0.59 |
| cg26599989 | chr11:1297087 | *TOLLIP* | rs5743936 | 0.26 | chr11:1316431 | G | 0.42 | 0.05 | 1.31E-16 | meQTL EPIC | A | G | -0.59 | 0.56 |
| cg26599989 | chr11:1297087 | *TOLLIP* | rs5743899 | 0.34 | chr11:1323564 | T | 0.21 | 0.04 | 4.06E-08 | meQTL EPIC | T | C | 0.42 | 0.68 |
| cg26599989 | chr11:1297087 | *TOLLIP* | rs187976925 | 0.03 | chr11:1420541 | G | -0.46 | 0.10 | 2.80E-06 | meQTL EPIC | NA | NA | NA | NA |
| cg19719391 | chr4:26789915 | *Intergenic* | rs114362105 | 0.006 | chr4:26573525 | T | -0.70 | 0.05 | 5.98E-49 | GoDMC | T | C | -1.43 | 0.15 |
| cg19719391 | chr4:26789915 | *Intergenic* | rs17639483 | 0.34 | chr4:26422507 | A | 0.16 | 0.03 | 1.14E-06 | meQTL EPIC | A | G | -2.10 | **3.61E-02** |

| | | | | | | | | | | | | | | |
|---|---|---|---|---|---|---|---|---|---|---|---|---|---|---|
| cg19719391 | chr4:26789915 | *Intergenic* | rs12646334 | 0.47 | chr4:26824285 | A | -0.25 | 0.05 | 5.38E-07 | meQTL EPIC | A | G | -0.76 | 0.45 |
| cg09822192 | chr14:24801191 | *ADCY4* | rs112679565 | 0.05 | chr14:24661174 | T | -0.49 | 0.01 | <5E-324 | GoDMC | T | C | 0.87 | 0.38 |
| cg09822192 | chr14:24801191 | *ADCY4* | rs2255591 | 0.35 | chr14:24901844 | A | -0.14 | 0.01 | 3.24E-47 | GoDMC | A | G | 0.87 | 0.38 |
| cg09822192 | chr14:24801191 | *ADCY4* | rs2243891 | 0.15 | chr14:24846757 | A | 0.47 | 0.02 | 3.13E-182 | GoDMC | A | G | 0.06 | 0.95 |
| cg09822192 | chr14:24801191 | *ADCY4* | rs17101367 | 0.33 | chr14:24587667 | T | -0.35 | 0.05 | 1.84E-13 | meQTL EPIC | T | C | 0.21 | 0.83 |
| cg09822192 | chr14:24801191 | *ADCY4* | chr14:24661633 | NA | chr14:24661633 | G | 0.42 | 0.08 | 2.90E-08 | meQTL EPIC | NA | NA | NA | NA |
| cg09822192 | chr14:24801191 | *ADCY4* | rs60283052 | 0.24 | chr14:24752137 | T | -0.70 | 0.06 | 9.41E-30 | meQTL EPIC | NA | NA | NA | NA |
| cg09822192 | chr14:24801191 | *ADCY4* | rs10483285 | 0.07 | chr14:24799358 | A | 0.84 | 0.08 | 4.43E-27 | meQTL EPIC | A | C | -3.56 | **3.73E-04** |
| cg09822192 | chr14:24801191 | *ADCY4* | rs2042052383 | <0.001 | chr14:24801443 | C | -0.67 | 0.04 | 3.97E-58 | meQTL EPIC | NA | NA | NA | NA |
| cg09822192 | chr14:24801191 | *ADCY4* | rs11621792 | 0.23 | chr14:24871926 | T | 0.38 | 0.04 | 5.87E-23 | meQTL EPIC | T | C | -0.48 | 0.63 |
| cg09822192 | chr14:24801191 | *ADCY4* | rs200457643 | <0.001 | chr14:25188870 | ATG | 0.15 | 0.03 | 3.18E-06 | meQTL EPIC | NA | NA | NA | NA |
| cg25691167 | chr7:19184961 | *FERD3L* | rs12700019 | 0.21 | chr7:19186886 | C | 0.18 | 0.01 | 3.15E-40 | GoDMC | C | G | -1.55 | 0.12 |
| cg11256214 | chr12:110211642 | *MGC14436* | rs11614658 | 0.18 | chr12:110082320 | C | -0.41 | 0.08 | 3.98E-07 | meQTL EPIC | T | C | 0.01 | 0.9903 |
| cg00774777 | chr11:76478902 | *RP11-21L23.4* | rs12794131 | 0.14 | chr11:76456739 | T | -0.37 | 0.04 | 1.44E-18 | meQTL EPIC | T | C | -1.05 | 0.2921 |
| cg00774777 | chr11:76478902 | *RP11-21L23.4* | rs60651758 | 0.096 | chr11:76469378 | A | 0.50 | 0.11 | 1.32E-05 | meQTL EPIC | A | G | -0.32 | 0.7519 |
| cg21566642 | chr2:233284661 | *Intergenic* | chr2:233221796 | 0.11 | chr2:233221796 | delT | -0.20 | 0.02 | <5E-183 | GoDMC | NA | NA | NA | NA |

Only significant ($p$ < 1E-08 for GoDMC and $p$ < 2.21E-04 for meQTL EPIC database) methylation quantitative trait loci (meQTL) were shown. All meQTLs are cis. No trans-meQTLs were identified. GWAS results are obtained from the Freeze 3 GWAS from Psychiatric Genomics Consortium PTSD Workgroup (PGC-PTSD)[73]. MAF column shows minor allele frequency from 1000Genomes phase 3 Global population. A1 indicates effect (tested) allele and A2 indicates alternative (reference) allele. D in A1 column indicates deletion. Beta: Linear regression beta coefficient. SD: Standard deviation. NA indicates that SNP was not available in the PGC-PTSD Freeze 3 GWAS summary statistics[73]. Significant GWAS results ($p$ < 0.05) are shown in bold. Positions are based on hg19.

**eTable 10.** Genetic associations of the identified PTSD-associated genes from gene-based tests.

| **Gene** | **Chr** | **Start** | **Stop** | **NSNPs** | **z** | ***p*-value** |
|---|---|---|---|---|---|---|
| *AHRR* | 5 | 304,291 | 438,406 | 390 | 2.70 | **3.48E-03** |
| *BANP* | 16 | 87,982,850 | 88,110,924 | 542 | -0.18 | 0.57 |
| *GFI1* | 1 | 92,940,319 | 92,952,433 | 20 | 0.99 | 0.16 |
| *TOLLIP* | 11 | 1,295,601 | 1,330,884 | 108 | -0.15 | 0.56 |
| *CHD5* | 1 | 6,161,853 | 6,240,183 | 252 | -0.61 | 0.73 |
| *CDC42BPB* | 14 | 103,398,716 | 103,523,799 | 283 | 3.20 | **6.94E-04** |
| *ADCY4* | 14 | 24,787,555 | 24,804,299 | 50 | 0.55 | 0.29 |
| *FERD3L* | NA | NA | NA | NA | NA | NA |
| *MGC14436* | NA | NA | NA | NA | NA | NA |
| *BCL11B* | 14 | 99,635,624 | 99,737,861 | 241 | 3.87 | **5.40E-05** |
| *RP11-21L23.4* | NA | NA | NA | NA | NA | NA |

Gene based test results are obtained from the Freeze 3 GWAS from PGC-PTSD[73]. Chr: chromosome. Start: Gene start position. Stop: Gene stop position. NSNPs: number of SNPs annotated to gene and used in the analysis. Significant results ($p$ < 0.05) are shown in bold.
